# Early transfusion of a large cohort of COVID-19 patients with high titer anti-SARS-CoV-2 spike protein IgG convalescent plasma confirms a signal of significantly decreased mortality

**DOI:** 10.1101/2020.10.02.20206029

**Authors:** Eric Salazar, Paul A. Christensen, Edward A. Graviss, Duc T. Nguyen, Brian Castillo, Jian Chen, Bevin Valdez Lopez, Todd N. Eagar, Xin Yi, Picheng Zhao, John Rogers, Ahmed Shehabeldin, David Joseph, Faisal Masud, Christopher Leveque, Randall J. Olsen, David W. Bernard, Jimmy Gollihar, James M. Musser

## Abstract

Coronavirus disease 2019 (COVID-19) caused by severe acute respiratory syndrome coronavirus 2 remains a global threat with few proven efficacious treatments. Transfusion of convalescent plasma collected from donors who have recovered from COVID-19 disease has emerged as a promising therapy and has been granted emergency use authorization by the U.S. Food and Drug Administration (FDA). We recently reported results from interim analysis of a propensity-score matched study suggesting that early treatment of COVID-19 patients with convalescent plasma containing high titer anti-spike protein receptor binding domain (RBD) IgG significantly decreases mortality. We here present results from 60-day follow up of our cohort of 351 transfused hospitalized patients. Prospective determination of ELISA anti-RBD IgG titer facilitated selection and transfusion of the highest titer units available. Retrospective analysis by the Ortho VITROS IgG assay revealed a median signal/cutoff (S/C) ratio of 24.0 for transfused units, a value far exceeding the recently FDA-required cutoff of 12.0 for designation of high titer convalescent plasma. With respect to altering mortality, our analysis identified an optimal window of 44 hours post-hospitalization for transfusing COVID-19 patients with high titer convalescent plasma. In the aggregate, the analysis confirms and extends our previous preliminary finding that transfusion of COVID-19 patients soon after hospitalization with high titer anti-spike protein RBD IgG present in convalescent plasma significantly reduces mortality.

## INTRODUCTION

Coronavirus disease 2019 (COVID-19) caused by severe acute respiratory syndrome coronavirus 2 (SARS-CoV-2) has caused massive societal disruption and death globally. As of September 27, 2020, there have been more than 33 million COVID-19 cases causing in excess of 1,000,000 deaths worldwide.^1^ The United States has many areas where rising case rates continue to threaten multiple populations. Very few effective treatments exist (*https://www.covid19treatmentguidelines.nih.gov/*), although hundreds of registered clinical trials are ongoing, including several phase 3 vaccine trials (*https://www.nytimes.com/interactive/2020/science/coronavirus-vaccine-tracker.html*, last accessed September 24, 2020).

We and others have published safety and efficacy outcomes in patients who were transfused with COVID-19 convalescent plasma.^2-4^ Aggregated available evidence stimulated the U.S. Food and Drug Administration (FDA) in late August 2020 to grant Emergency Use Authorization (EUA) for COVID-19 convalescent plasma therapy (*https://www.fda.gov/media/141477/download*, last accessed September 24, 2020). In our previous study, interim analysis revealed that, relative to matched controls, patients transfused with convalescent plasma containing high titer anti-spike protein receptor binding domain (RBD) IgG within 72 hrs of hospital admission had significantly reduced mortality at 28 days post-transfusion.^3^

To further investigate these observations, and to address limitations inherent in an interim analysis, we here present results from 60-day follow up of our entire cohort of 351 transfused patients. The data confirm our previous findings that transfusion of patients soon after hospital admission with high titer anti-spike protein RBD IgG present in convalescent plasma significantly decreases mortality.

## MATERIALS AND METHODS

We analyzed data from patients cared for in all eight Houston Methodist hospitals from March 28, 2020, through September 14, 2020, with the approval of the Houston Methodist Research Institute ethics review board and with written informed consent of the patient or legally authorized representative. Details of the study, including inclusion and exclusion criteria, and criteria for the transfusion of multiple units have been described.^3^

### Convalescent plasma donors, antibody titer assessment, and donor unit selection

Detailed protocols for convalescent plasma collection and anti-spike protein titer assessment have been described.^*3, 5, 6*^ COVID-19 convalescent plasma units were selected for transfusion based on anti-spike ectodomain and RBD IgG ELISA titers available on donor units obtained from April 7, 2020 onward. We previously published that plasma with an anti-RBD IgG titer of ≥1:1350 corresponds to an ∼80% probability of a live virus *in vitro* neutralization titer of ≥1:160.^7^ This titer is the value initially recommended by the FDA for transfusing COVID-19 patients.^8^ To facilitate the need for increased donor unit assessment, we standardized our ELISA to four plasma dilutions: 1:50, 1:150, 1:450, and 1:1350. To select the highest titer unit available, ELISA results were ranked based on highest titer and, subsequently by highest optical density at dilution 1:50. Patients were transfused with the ABO-compatible convalescent plasma unit with the highest titer and highest optical density at dilution 1:50 available. Frozen serum samples were assessed retrospectively with the Ortho VITROS IgG assay (Raritan, NJ) according to manufacturer’s instructions.

### Statistical analysis

We analyzed patients who met a 60-day outcome defined as having outcome data available 60 days post-transfusion (cases) and 60 days post-hospitalization (controls). Control patients enrolled in other clinical trials were excluded from the analysis. Patients discharged before Day 60 were presumed to be on room air after discharge unless otherwise noted in the electronic medical record. Baseline characteristics for COVID-19 patients who met the 60-day outcome definition are shown in **Table S1**.

We conducted a one-to-many nearest neighbor propensity score matching analysis without replacement using an initial ratio of case:control=1:3 and caliper of ≤1 between patients having plasma transfusion (cases) versus patients who did not have plasma transfusion (controls). The primary matching criteria included age (categorical, <30, 30-39, 40-49, 50-59, 60-69, 70-79, ≥80), sex, BMI (+/- 30), diabetes, hypertension, chronic pulmonary disease, chronic kidney disease, hyperlipidemia, coronary disease, and baseline ventilation requirement within 48 hrs of admission, use of any steroid, azithromycin, hydroxychloroquine, remdesivir, ribavirin, and tocilizumab. A secondary propensity score matching was conducted based on the ventilation status at Day 0, defined as the day of transfusion for cases and the corresponding day in the hospitalization course for controls, using a case:control ratio of either 1:2 or 1:1 and caliper ≤1.^9^

The primary outcome (mortality within 60 days post-Day 0) was displayed by Kaplan-Meier curves. Differences between groups were compared with the log-rank test. Cox proportional hazards modeling (with clustered sandwich estimator option for the matched cluster in the propensity-matched cohorts) was performed to determine the characteristics associated with the overall mortality within 28 days and 60 days. Variables for the multivariable models were selected based on potential clinical relevance and using Stata’s Lasso technique with cross-validation.^10, 11^ Receiver operating characteristic (ROC) curve analysis with Youden index was used to identify the optimal time (in hours) from admission-to-transfusion of first unit that discriminates 60-day mortality in patients that received COVID-19 convalescent plasma.^12^

Generalized linear modeling (GLM) and multinomial logistic regression with a cluster variance estimator were also used to evaluate several exploratory endpoints. The evaluated covariates included supplemental oxygen requirements (room air, low-flow oxygen delivery, high-flow oxygen delivery, non-invasive positive pressure ventilation, mechanical ventilation, extracorporeal membrane oxygenation (ECMO), or death) at Day 7, Day 14, Day 28, and Day 60 post-transfusion; clinical improvement relative to Day 0; intensive care unit (ICU) stay requirement; ICU length of stay; mechanical ventilation requirement; length of mechanical ventilation requirement; length of supplemental oxygen requirement; and inflammatory marker levels (interleukin-6, C-reactive protein, ferritin, fibrinogen, D-dimer) at Day 7. Clinical improvement relative to Day 0 was defined as a 1 point improvement in ordinal scale [1, discharged (alive); 2, hospitalized, not requiring supplemental oxygen but requiring ongoing medical care (for COVID-19 or otherwise); 3, hospitalized, requiring low-flow supplemental oxygen; 4, hospitalized, on non-invasive ventilation or high-flow oxygen devices; 5, hospitalized and on invasive mechanical ventilation or ECMO; 6, death]. All analyses were performed with Stata version 16.1 (StataCorp LLC, College Station, TX, USA) or the R Statistical Computing environment (http://www.R-project.org/). A *p*-value of ≤0.05 was considered significant.

## RESULTS

### Study population and baseline characteristics

We had 5,297 hospitalized COVID-19 patients available for analysis, 353 of whom were transfused with COVID-19 convalescent plasma. Two of the 353 patients received plasma without a titer assessment prior to transfusion, and these patients were excluded from the overall analysis, resulting in a cohort of 351 transfused evaluable patients. Relative to non-transfused patients, transfused patients were significantly younger, predominantly male, predominantly Hispanic, had a higher BMI, lower rates of comorbidities (specifically, chronic pulmonary disease, chronic kidney disease, hyperlipidemia, and coronary disease, but not hypertension and diabetes), a higher requirement for supplemental oxygen, and higher inflammatory biomarker concentrations. D-dimer was significantly lower in the transfused cohort at baseline by 0.2 fibrinogen equivalent units. Use of steroids, azithromycin, remdesivir, and tocilizumab was more common among the transfused cohort (**Table S1**).

### Safety

Among 351 transfused patients included in the study, only seven (2.0%) had adverse events deemed related to plasma transfusion. Six events were classified as allergic transfusion reactions and five of these six were mild and included only a transient rash. One patient developed transient worsening of shortness of breath that resolved with diphenhydramine. One case of possible transfusion-associated circulatory overload occurred, with associated transient worsening of dyspnea that improved with furosemide. These two events were deemed to be significant adverse events. Thus, among the 351 transfused study patients, only two (0.6%) significant adverse events were deemed related to plasma transfusion.

### Factors associated with a higher risk of death in all hospitalized COVID-19 patients

Univariate and multivariate Cox proportional hazards modeling assessing factors associated with a higher risk of death within 60 days post-transfusion Day 0 was performed for all COVID-19 patients admitted to our eight hospitals during the study period for whom data were available (**Tables S2 and S3**). Factors associated with a higher risk of death in the multivariate analysis included age, male sex, diabetes, chronic kidney disease, worst ventilation status within 48 hrs of admission, and/or administration of any steroids or tocilizumab. Neither ABO blood type, race, nor ethnicity were associated with higher risk of death in the multivariate analysis. Importantly, the covariates that had a significant association with risk of death were included in the propensity score matching algorithm. We did not include baseline inflammatory concentrations in the multivariate analysis and in the propensity score matching algorithm because of the high proportion of missing data.

### COVID-19 convalescent plasma and retrospective analysis of Ortho VITROS IgG test data

Most transfused patients (278/351; 79%) received only one ∼300 mL unit of COVID-19 convalescent plasma. The great majority of patients received an initial or sole unit of convalescent plasma with anti-RBD IgG titer of ≥1:1350 (321/351; 91%); 24 patients received an initial or sole unit of convalescent plasma with an anti-RBD IgG titer >1:150 but <1:1350; six patients received an initial or sole unit of convalescent plasma with anti-RBD IgG titer of <1:150. For patients who received a second unit of convalescent plasma, 71 (71/75; 95%) received a second unit with an anti-RBD IgG titer ≥1:1350, and four (4/75; 5%) patients received a second unit with an anti-RBD IgG titer >1:150 but <1:1350.

The FDA issued an EUA for convalescent plasma transfusion of COVID-19 patients on August 23, 2020. The agency’s guidance is to use convalescent plasma units with an S/C level of >12, as defined by the Ortho VITROS IgG test (*https://www.fda.gov/media/141477/download*, last accessed September 24, 2020). For 278 of the 351 (79%) initial plasma units transfused, a sample was available for retrospective assessment of anti-SARS-CoV-2 IgG titer by the Ortho VITROS IgG test. The median IgG signal/cutoff (S/C) ratio was 24.0 (range=0.01-35) and only seven units (3%) had a corresponding S/C ratio of <12. In addition, we found a very strong positive correlation between the ELISA anti-RBD IgG optical density at dilution 1:50 and the Ortho VITROS IgG test for 1,142 samples with parallel assessment (R=0.88; *P*<0.001). The distribution of Ortho VITROS IgG S/C ratios and anti-RBD IgG ELISA optical density for transfused plasma units confirms that high anti-spike protein IgG titer units were being given to the enrolled COVID-19 patients (**Figure 1**).

**Figure 1.**
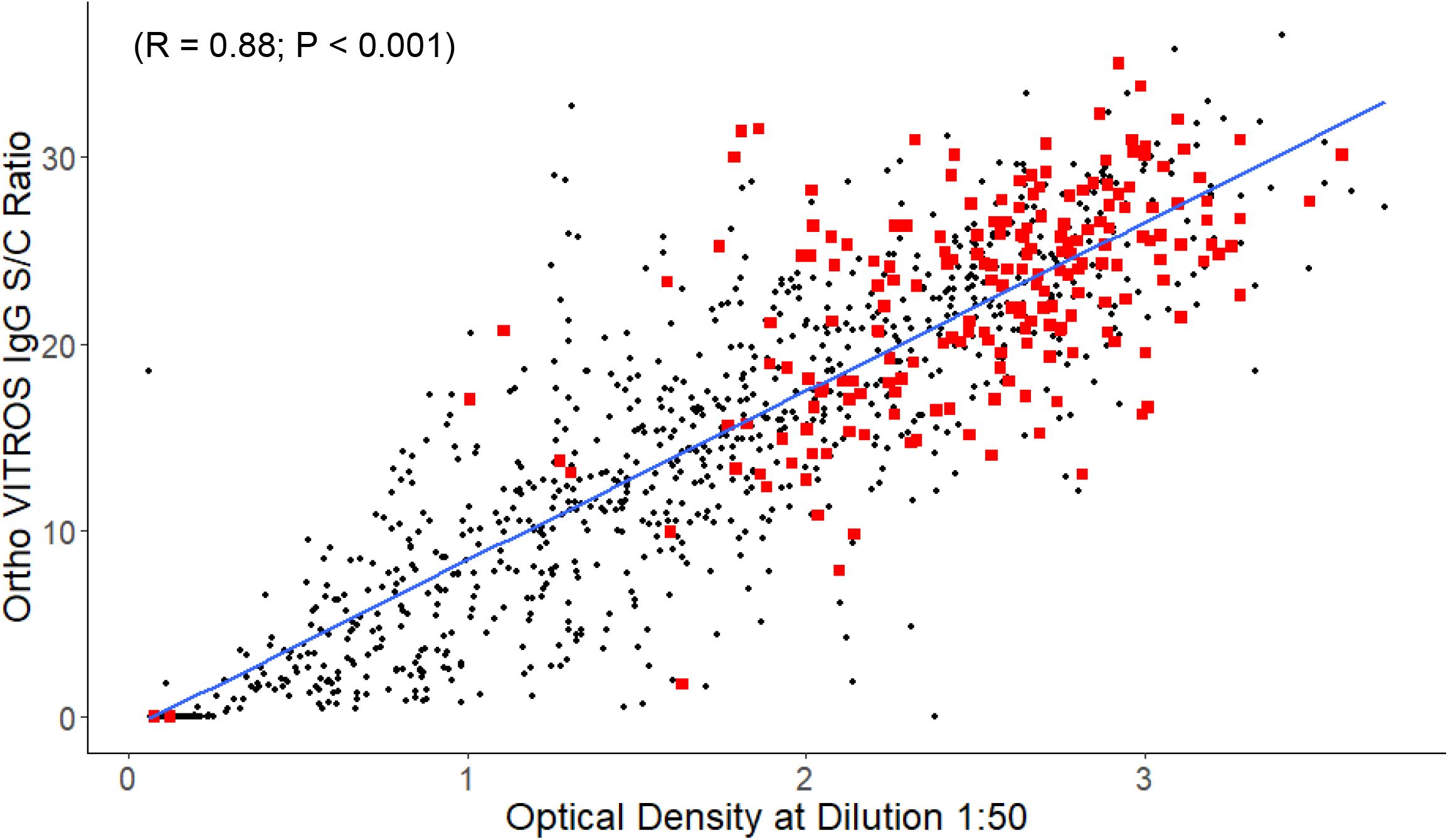
Ortho VITROS IgG signal/cutoff (S/C) ratio versus optical density at dilution 1:50 for serum samples for all convalescent plasma collections and for which parallel testing data was available through September 27, 2020. The blue line is the linear regression line of best fit. Positive linear correlation was significant (R=0.88; *P*<0.001). Red squares denote units transfused in the study. Black circles denote samples for all other units collected and not transfused during the study. Many of these units (black circles) were deferred due to the presence of donor HLA antibodies or positive donor SARS-CoV-2 nasopharyngeal swab at the time of donation.

### Outcomes

Propensity score matching yielded a study population of 341 transfused patients and 594 matched controls, which were balanced across all matching criteria (**Figure 2 and Table S4**). Kaplan-Meier curves showed significantly decreased mortality within 60 days post-Day 0 in the transfused cohort relative to propensity score-matched controls (*P*=0.02) (data not shown). Statistical significance increased to *P*=0.003 when the matching algorithm and analysis were restricted to patients transfused with plasma with an anti-RBD IgG titer of ≥1:1350 (**Figure 3**). Mortality was not significantly different within 60 days post-Day 0 between cases and controls in patients who were intubated at Day 0 or in patients who were transfused more than 72 hrs after admission, even when the analysis was restricted to patients who received plasma with a high titer anti-RBD IgG. There was no significant difference in mortality between cases and controls when the analysis was restricted to patients who received plasma with an anti-RBD IgG titer of <1:1350. In contrast, mortality was significantly decreased in patients who received plasma with an anti-RBD IgG titer of ≥1:1350 within 72 hrs of admission (**Figure 4**). Point estimates of the outcomes when the analysis was restricted to transfusion of high titer plasma confirm these findings (**Table 1**).

**Figure 2.**
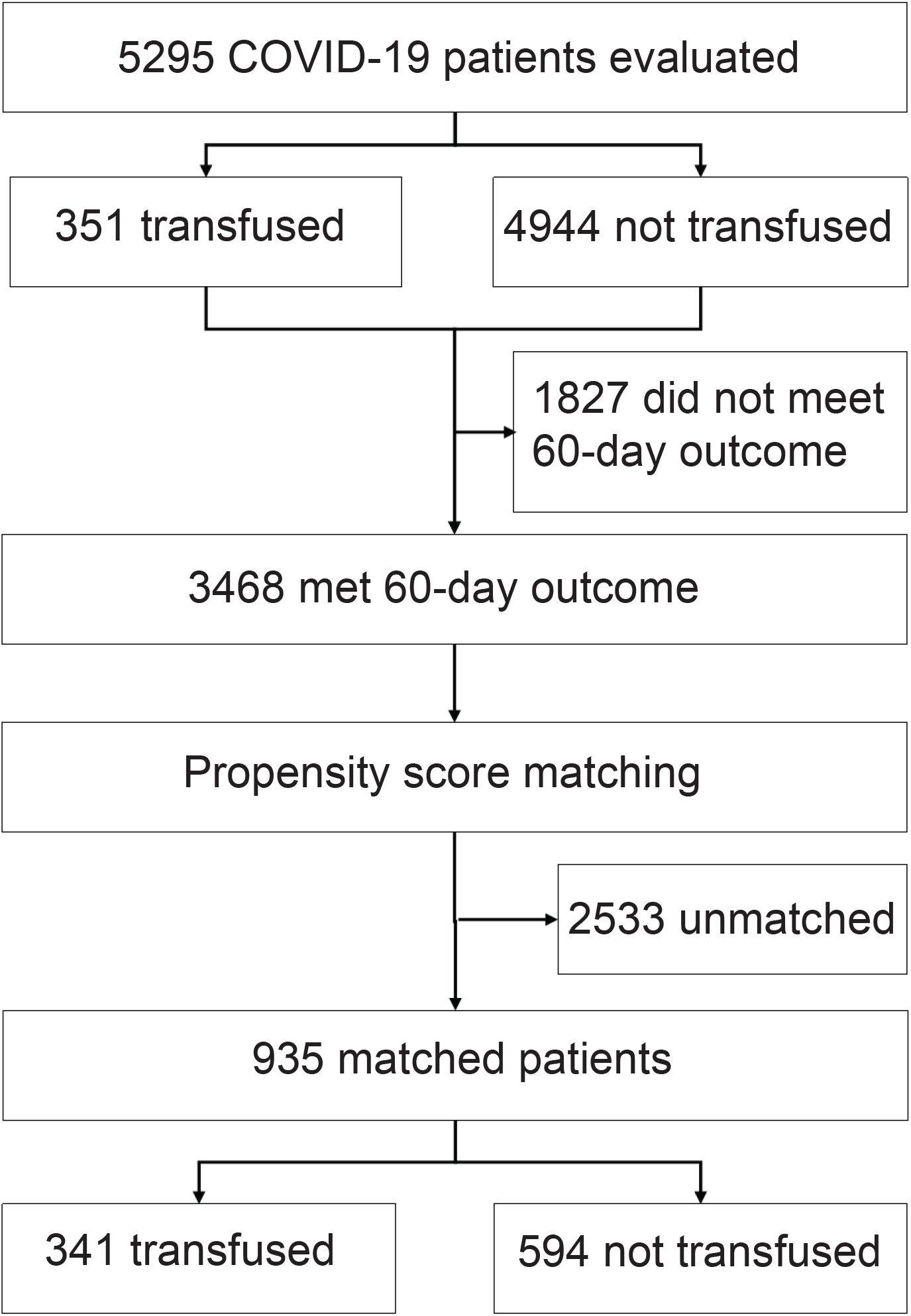
Flowchart of the study population. Propensity score matching was based on patient age (categorical, per 10 years); sex; BMI (categorical, +/- 30); presence of diabetes, hypertension, chronic pulmonary disease, chronic kidney disease, hyperlipidemia and/or coronary disease; baseline ventilation status within 48 hrs of admission (room air, supplemental oxygen, and mechanical ventilation); and use of any steroid, azithromycin, hydroxychloroquine, remdesivir, ribavirin, and tocilizumab. After establishing the first propensity score-matched cohort and obtaining Day 0 for controls, a second match was run between cases and controls based on the ventilation status at Day 0.

**Figure 3.**
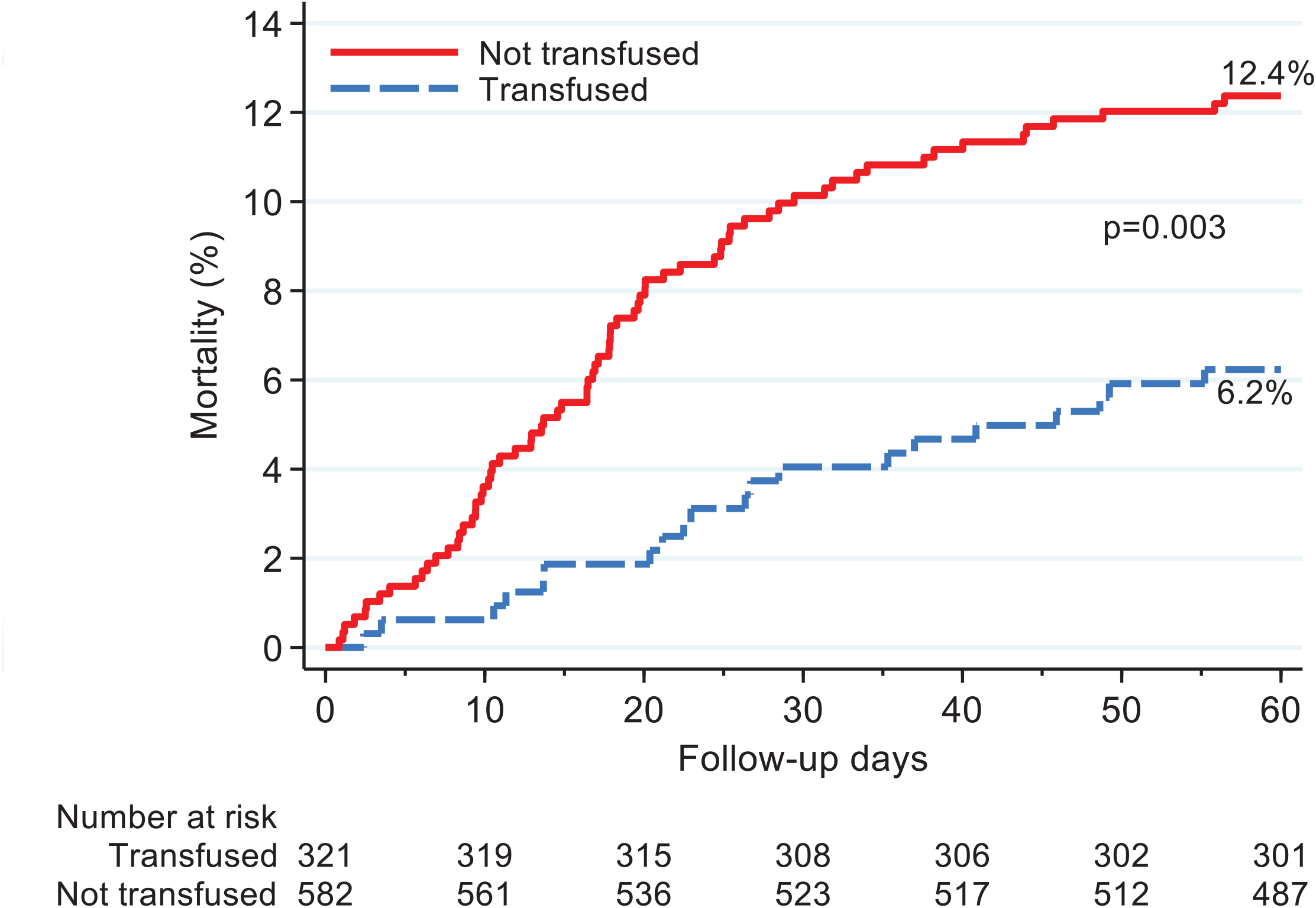
Kaplan-Meier curves for mortality within 60 days post-Day 0 for all patients who received plasma with an anti-RBD IgG titer ≥1:1350 regardless of time from admission (blue) propensity score-matched to controls (red).

**Figure 4.**
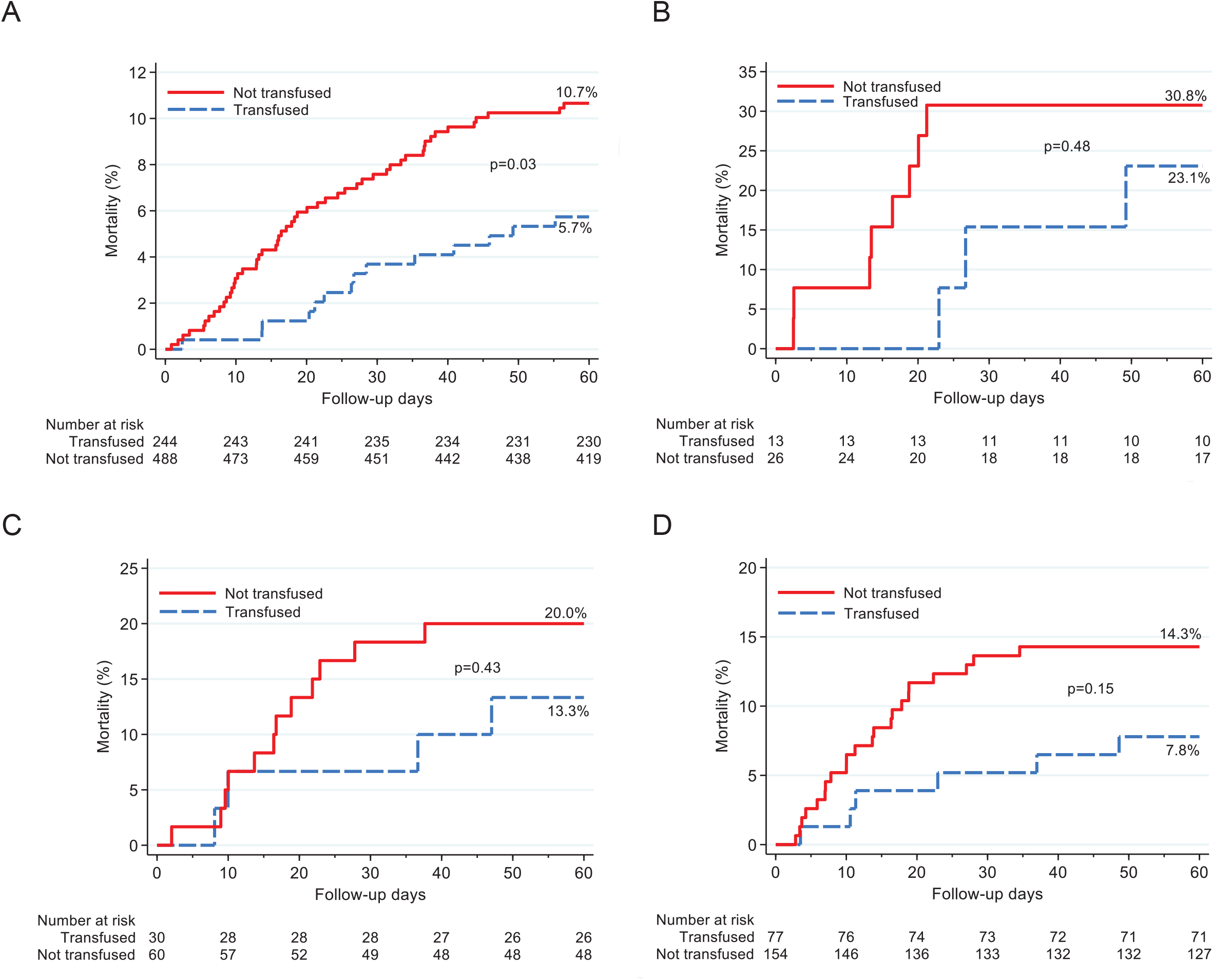
Kaplan-Meier curves for mortality within 60 days post-Day 0 for different cohorts of propensity-score matched patients and controls. A) Patients transfused with plasma with an anti-RBD IgG titer ≥1:1350 and transfused within 72 hrs of admission (blue) propensity score-matched to control patients (red). B) Patients transfused with plasma with an anti-RBD IgG titer ≥1:1350 and intubated at Day 0 (blue) propensity score-matched to control patients intubated at Day 0 (red). C) Patients transfused with plasma with an anti-RBD IgG titer <1:1350 (blue) propensity score-matched to control patients (red). D) Patients transfused with plasma with an anti-RBD IgG titer ≥1:1350 and transfused greater than 72 hrs after admission (blue) propensity score-matched to control patients (red).

**Table 1.**
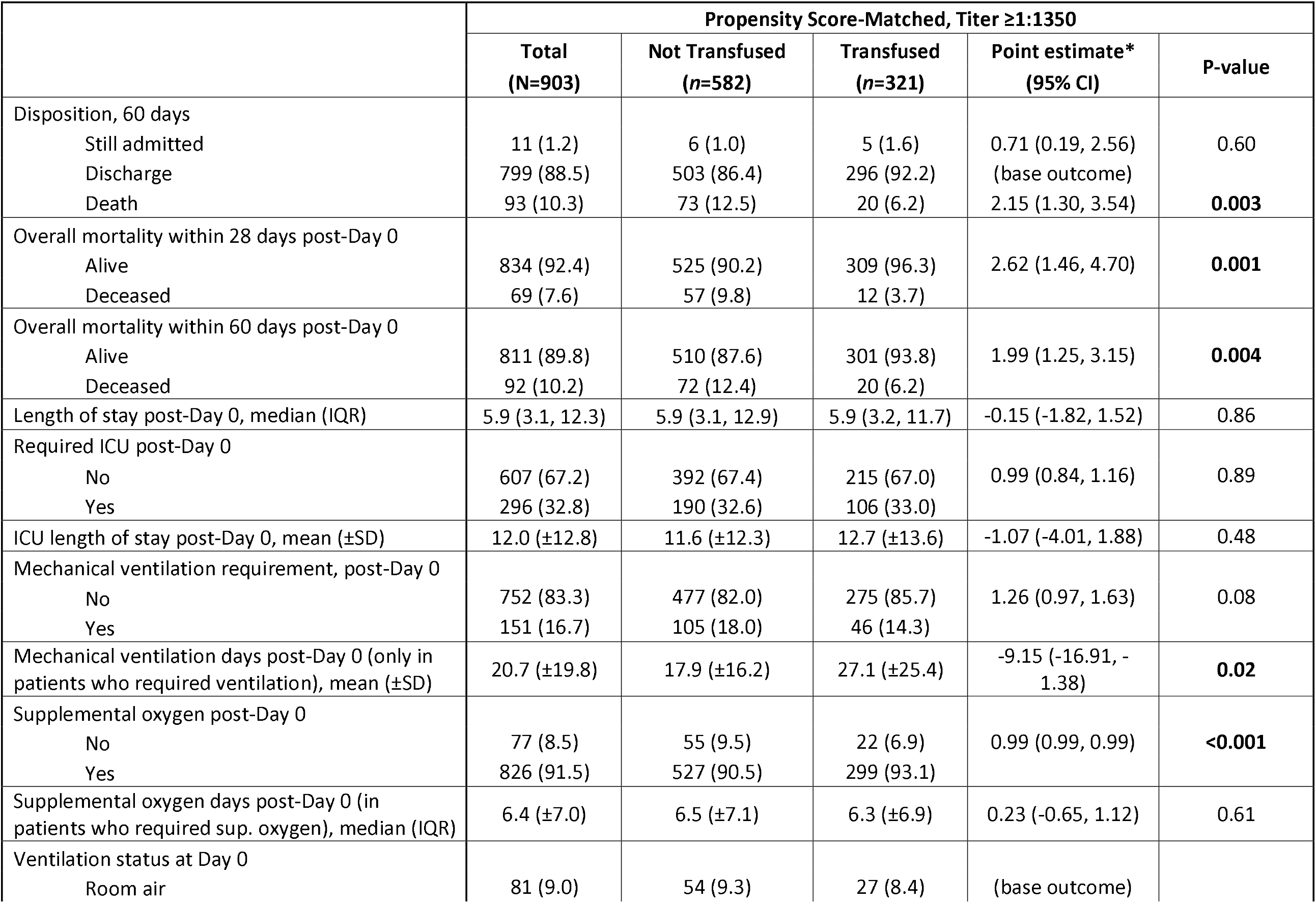
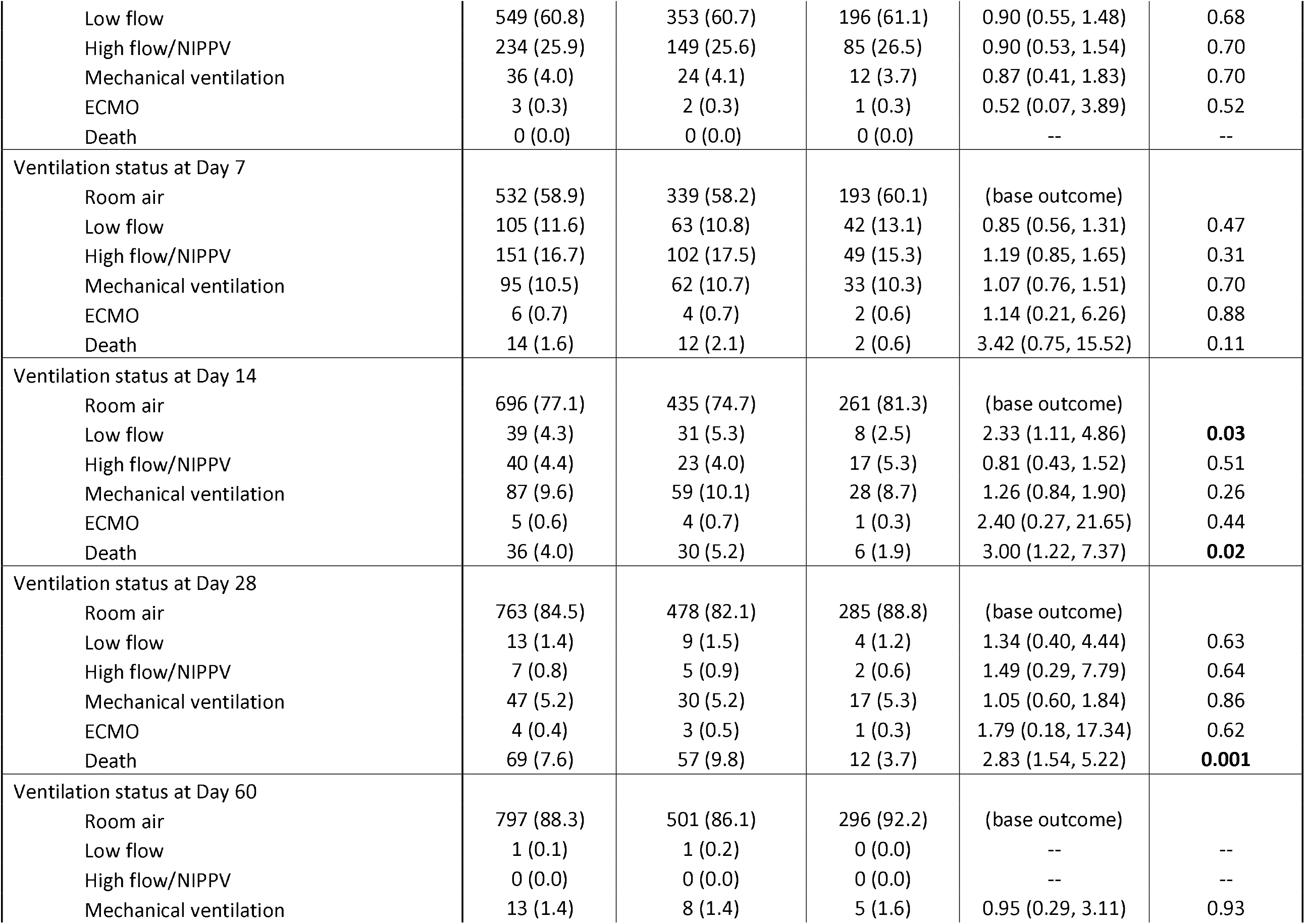
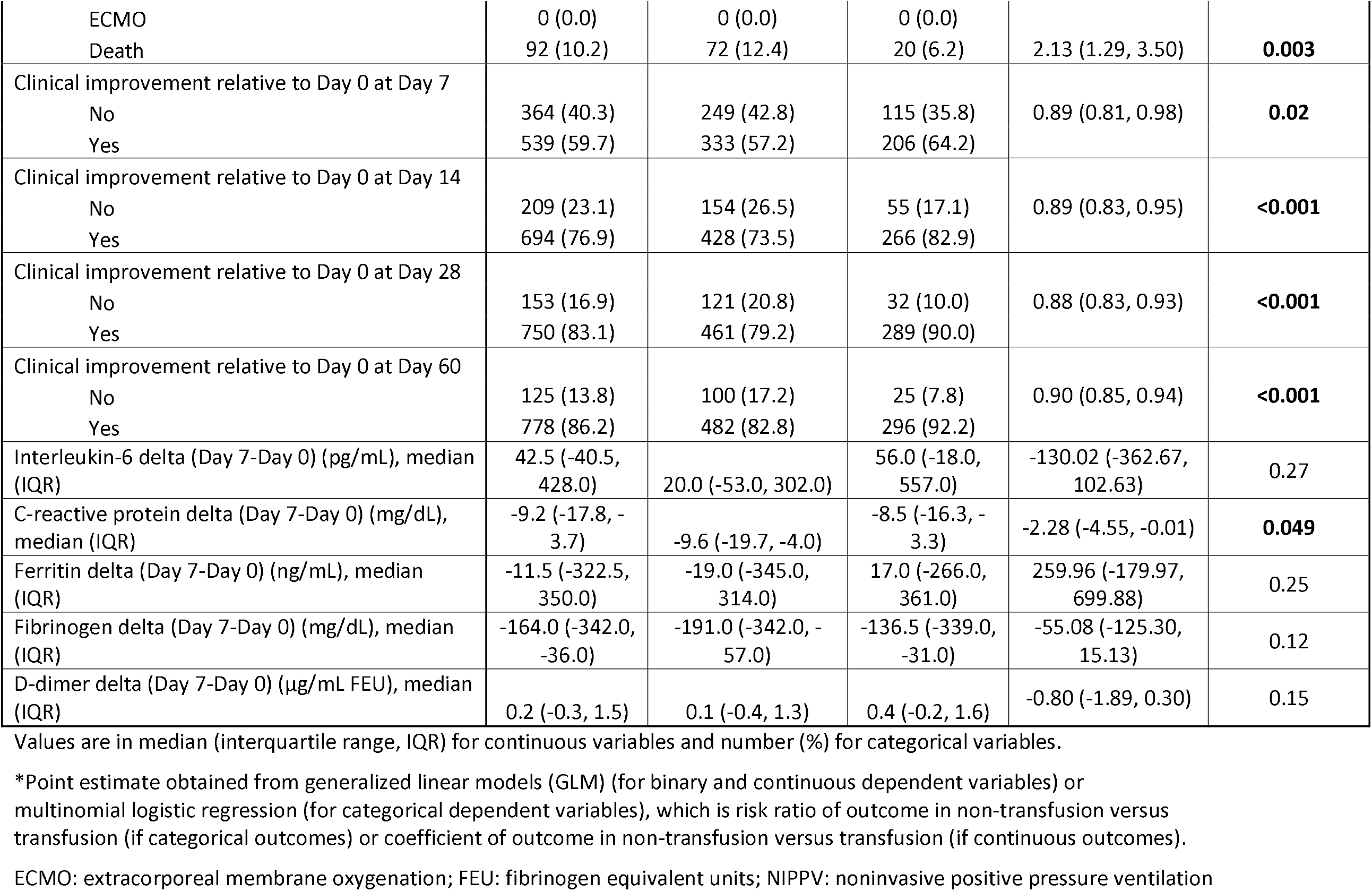
Outcome Summary

Consistent with these observations, the unadjusted HR and adjusted HR in the univariate and multivariate Cox proportional hazards models for mortality within 60 days was significant when the analysis was restricted to patients who received plasma with an anti-RBD IgG titer of ≥1:1350 (**Table 2**). Due to small sample sizes, multivariate analysis could not be performed for patients who received plasma with a titer ≥1:1350 and were intubated at Day 0, or who were transfused more than 72 hrs after hospitalization. In these two cohorts, the unadjusted HR in univariate analyses for mortality within 60 days post-Day 0 was not significant (HR=1.61 for controls; *P*=0.44 and HR=1.93 for controls; *P*=0.16, respectively). Similarly, the unadjusted HR for mortality within 60 days in the analysis restricted to patients who received plasma with a titer <1:1350 was not significant (HR=1.57 for controls, *P*=0.36). However, the unadjusted HR for mortality within 60 days was significant (HR=1.93 for controls, *P*=0.02) when the analysis was restricted to patients who received plasma with a titer ≥1:1350 within 72 hrs of hospital admission. For this cohort, the adjusted HR for mortality within 60 days was significant when assessed for a 28-day outcome (aHR=2.09 for controls; *P*=0.047) and approached significance when assessed for a 60-day outcome (aHR=1.82 for controls; *P*=0.051).

**Table 2.** Univariate and Multivariate Cox Regression, Overall Mortality within 28 and 60 days, Controls Matched to Cases that Received Plasma with Titer ≥1:1350

| Univariate | Within 60 days |  |  |  |
| --- | --- | --- | --- | --- |
|  | Alive<br>(n=811) | Deceased<br>(n=92) | Unadjusted HR<br>(95% CI) | P-value |
| Convalescent plasma transfusion |  |  |  |  |
| Transfused | 301 (37.1) | 20 (21.7) | (reference) |  |
| Not Transfused | 510 (62.9) | 72 (78.3) | 1.07 (1.05, 1.09) | **<0.001** |
| Age (years), median (IQR) | 54.0 (44.0, 62.0) | 65.0 (59.0, 76.0) | 1.07 (1.05, 1.09) | **<0.001** |
| Age (years) |  |  |  |  |
| <30 | 33 (4.1) | 3 (3.3) | 3.65 (0.66, 20.33) | 0.14 |
| 30-39 | 111 (13.7) | 3 (3.3) | 1.14 (0.26, 5.02) | 0.87 |
| 40-49 | 171 (21.1) | 4 (4.3) | (reference) |  |
| 50-59 | 226 (27.9) | 16 (17.4) | 2.94 (0.97, 8.91) | 0.06 |
| 60-69 | 182 (22.4) | 30 (32.6) | 6.54 (2.29, 18.72) | **<0.001** |
| 70-79 | 69 (8.5) | 20 (21.7) | 10.85 (3.66, 32.19) | **<0.001** |
| $\geq 80$ | 19 (2.3) | 16 (17.4) | 29.06 (8.94, 94.50) | **<0.001** |
| Sex |  |  |  |  |
| Female | 362 (44.6) | 33 (35.9) | (reference) |  |
| Male | 449 (55.4) | 59 (64.1) | 1.41 (0.92, 2.16) | 0.12 |
| Race |  |  |  |  |
| White | 530 (65.4) | 65 (70.7) | (reference) |  |
| Black | 185 (22.8) | 16 (17.4) | 0.73 (0.43, 1.24) | 0.25 |
| Asian | 41 (5.1) | 5 (5.4) | 1.02 (0.40, 2.58) | 0.97 |
| Other | 25 (3.1) | 5 (5.4) | 1.53 (0.67, 3.49) | 0.31 |
| Unknown | 30 (3.7) | 1 (1.1) | 0.28 (0.04, 2.07) | 0.21 |
| Ethnicity |  |  |  |  |
| Non-Hispanic | 399 (49.2) | 48 (52.2) | (reference) |  |
| Hispanic | 406 (50.1) | 42 (45.7) | 0.86 (0.58, 1.26) | 0.43 |
| Unknown | 6 (0.7) | 2 (2.2) | 2.52 (0.61, 10.53) | 0.20 |
| Body mass index (kg/m<sup>2</sup>), median (IQR) | 31.8 (27.8, 37.7) | 30.1 (26.7, 34.7) | 0.97 (0.94, 1.01) | 0.10 |
| Body mass index (kg/m2) |  |  |  |  |
| <18.5 | 2 (0.2) | 0 (0.0) | -- | -- |
| 18.5-24.9 | 88 (10.9) | 11 (12.0) | (reference) |  |
| 25-29.9 | 220 (27.1) | 32 (34.8) | 1.17 (0.60, 2.28) | 0.65 |
| ≥30 | 501 (61.8) | 49 (53.3) | 0.79 (0.41, 1.54) | **0.49** |
| Body mass index ≥30 (kg/m2) |  |  |  |  |
| <30 | 310 (38.2) | 43 (46.7) | (reference) |  |
| ≥30 | 501 (61.8) | 49 (53.3) | 0.71 (0.46, 1.10) | 0.12 |
| Hypertension |  |  |  |  |
| No | 396 (48.8) | 35 (38.0) | (reference) |  |
| Yes | 415 (51.2) | 57 (62.0) | 1.51 (0.98, 2.31) | 0.06 |
| Diabetes |  |  |  |  |
| No | 488 (60.2) | 39 (42.4) | (reference) |  |
| Yes | 323 (39.8) | 53 (57.6) | 1.96 (1.30, 2.96) | **0.001** |
| Chronic pulmonary disease |  |  |  |  |
| No | 721 (88.9) | 74 (80.4) | (reference) |  |
| Yes | 90 (11.1) | 18 (19.6) | 1.85 (1.12, 3.08) | **0.02** |
| Chronic kidney disease |  |  |  |  |
| No | 708 (87.3) | 64 (69.6) | (reference) |  |
| Yes | 103 (12.7) | 28 (30.4) | 2.80 (1.79, 4.38) | **<0.001** |
| Hyperlipidemia |  |  |  |  |
| No | 541 (66.7) | 48 (52.2) | (reference) |  |
| Yes | 270 (33.3) | 44 (47.8) | 1.79 (1.16, 2.76) | **0.01** |
| Coronary disease |  |  |  |  |
| No | 753 (92.8) | 67 (72.8) | (reference) |  |
| Yes | 58 (7.2) | 25 (27.2) | 4.46 (2.77, 7.16) | **<0.001** |
| Baseline outcome group |  |  |  |  |
| Room air | 38 (4.7) | 5 (5.4) | (reference) |  |
| Supplemental oxygen | 750 (92.5) | 71 (77.2) | 0.74 (0.30, 1.79) | 0.50 |
| Mechanical ventilation | 23 (2.8) | 16 (17.4) | 4.33 (1.56, 12.04) | **0.01** |
| Ventilation status at Day 0 |  |  |  |  |
| Room air | 78 (9.6) | 3 (3.3) | (reference) |  |
| Low flow | 516 (63.6) | 33 (35.9) | 1.66 (0.53, 5.15) | 0.38 |
| High flow/NIPPV | 194 (23.9) | 40 (43.5) | 4.99 (1.63, 15.25) | **0.01** |
| Mechanical ventilation | 20 (2.5) | 16 (17.4) | 15.69 (4.74, 52.02) | **<0.001** |
| ECMO | 3 (0.4) | 0 (0.0) | -- | -- |
| ABO blood group |  |  |  |  |
| A | 204 (30.6) | 25 (29.4) | 0.91 (0.58, 1.42) | 0.67 |
| B | 93 (14.0) | 9 (10.6) | 0.74 (0.36, 1.52) | 0.42 |
| AB | 13 (2.0) | 3 (3.5) | 1.59 (0.52, 4.84) | 0.42 |
| O | 356 (53.5) | 48 (56.5) | (reference) |  |
| Rh blood group |  |  |  |  |
| Negative | 54 (8.1) | 11 (12.9) | (reference) |  |
| Positive | 612 (91.9) | 74 (87.1) | 0.62 (0.34, 1.15) | 0.13 |
| Interleukin-6 at baseline, (pg/mL), median (IQR) (**n=604**) | 51.0 (23.0, 114.0) | 106.0 (35.0, 309.0) | 1.001 (1.00, 1.001) | **<0.001** |
| Interleukin-6 delta (Day 7-baseline), (pg/mL), median (IQR) (**n=236**) | 26.0 (-51.0, 232.0) | 496.0 (43.0, 966.0) | 1.0004 (1.00, 1.001) | **0.01** |
| C-reactive protein at baseline, (mg/dL), median (IQR) (**n=725**) | 9.7 (5.3, 16.4) | 10.2 (6.2, 15.2) | 0.99 (0.97, 1.02) | 0.65 |
| C-reactive protein delta (Day 7-baseline), (mg/dL), median (IQR) (**n=403**) | -9.5 (-18.3, -3.6) | -8.0 (-14.4, -4.3) | 1.01 (0.99, 1.03) | 0.30 |
| Ferritin at baseline, (ng/mL), median (IQR) (**n=726**) | 809.5 (427.0, 1565.0) | 1160.5 (543.5, 1896.0) | 1.00 (1.00, 1.00) | 0.052 |
| Ferritin delta (Day 7-baseline), (ng/mL), median (IQR) (**n=396**) | -20.5 (-351.0, 308.0) | 141.5 (-272.0, 591.0) | 1.0002 (1.00, 1.0003) | **0.03** |
| Fibrinogen at baseline, (mg/dL), median (IQR) (**n=534**) | 658.0 (562.0, 749.0) | 617.0 (526.0, 720.0) | 0.99 (0.99, 1.00) | **0.01** |
| Fibrinogen delta (Day 7-baseline), (mg/dL), median (IQR) (**n=155**) | -178.0 (-347.5, -48.5) | -121.0 (-246.0, 49.0) | 1.00 (1.00, 1.00) | 0.37 |
| D-dimer at baseline, (µg/mL FEU), median (IQR) (**n=733**) | 0.9 (0.6, 1.5) | 1.2 (0.7, 3.1) | 1.12 (1.07, 1.16) | **<0.001** |
| D-dimer delta (Day 7-baseline), (µg/mL FEU), median (IQR) (**n=394**) | 0.1 (-0.3, 1.0) | 1.3 (0.1, 11.5) | 1.08 (1.04, 1.12) | **<0.001** |
| Concomitant medication |  |  |  |  |
| Any steroids | 576 (71.0) | 87 (94.6) | 6.64 (2.72, 16.19) | **<0.001** |
| Dexamethasone | 395 (48.7) | 46 (50.0) | 1.04 (0.67, 1.60) | 0.87 |
| Hydrocortisone | 28 (3.5) | 33 (35.9) | 9.42 (6.12, 14.50) | **<0.001** |
| Methylprednisolone | 297 (36.6) | 67 (72.8) | 4.21 (2.65, 6.67) | **<0.001** |
| Prednisone | 109 (13.4) | 23 (25.0) | 1.96 (1.26, 3.04) | 0.00 |
| Azithromycin | 596 (73.5) | 68 (73.9) | 1.01 (0.63, 1.60) | 0.98 |
| Hydroxychloroquine | 80 (9.9) | 12 (13.0) | 1.37 (0.77, 2.42) | 0.28 |
| Lopinavir/ritonavir | 7 (0.9) | 2 (2.2) | 2.32 (0.57, 9.46) | 0.24 |
| Remdesivir | 306 (37.7) | 34 (37.0) | 0.95 (0.61, 1.49) | 0.84 |
| Ribavirin | 27 (3.3) | 4 (4.3) | 1.33 (0.48, 3.66) | 0.58 |
| Tocilizumab | 358 (44.1) | 73 (79.3) | 4.49 (2.70, 7.48) | **<0.001** |
| **Multivariate** | **Within 28 days** |  | **Within 60 days** |  |
|  | **Adjusted HR**<br>**(95% CI)** | **P-value** | **Adjusted HR**<br>**(95% CI)** | **P-value** |
| Convalescent plasma transfusion |  |  |  |  |
| Transfused | (reference) |  | (reference) |  |
| Not Transfused | 1.94 (1.05, 3.58) | **0.04** | 1.64 (1.00, 2.69) | **0.049** |
| Age (years) | 1.06 (1.04, 1.09) | **<0.001** | 1.09 (1.05, 1.13) | **<0.001** |
| Diabetes | 1.69 (1.01, 2.84) | **0.046** | 1.57 (1.02, 2.43) | **0.04** |
| Chronic kidney disease | 1.44 (0.79, 2.60) | 0.23 | 1.41 (0.83, 2.40) | 0.20 |
| Ventilation status at Day 0 |  |  |  |  |
| Room air | (reference) |  |  |  |
| Low flow | 3.42 (0.51, 23.12) | 0.21 | 1.46 (0.49, 4.34) | 0.50 |
| High flow/NIPPV | 5.14 (0.75, 35.13) | 0.10 | 2.71 (0.96, 7.64) | 0.06 |
| Mechanical ventilation | 12.99 (1.80, 93.62) | **0.01** | 5.68 (1.91, 16.90) | **0.002** |
| ECMO | -- | -- | -- | -- |
| Any steroids | 1.11 (1.03, 1.21) | **0.01** | 1.06 (1.00, 1.13) | 0.06 |
| Tocilizumab | 1.06 (1.01, 1.11) | **0.01** | 1.04 (1.01, 1.06) | **0.01** |
| C-statistic | **C-statistic = 0.87** | -- | **C-statistic = 0.81** | -- |

Values are in median (interquartile range, IQR) for continuous variables and number (%) for categorical variables.
Steroids and tocilizumab were treated as time-varying covariates in the multivariate model.
CI: confidence interval; HR: hazard ratio; ECMO: extracorporeal membrane oxygenation; FEU: fibrinogen equivalent units; NIPPV: noninvasive positive pressure ventilation

We sought to identify the optimal window after hospitalization within which transfusion of convalescent plasma was most useful with respect to altering mortality. ROC curve analysis with Youden index revealed an optimal cut point of transfusion within 44 hrs of hospital admission for discriminating mortality within 60 days post-transfusion in all patients transfused with COVID-19 convalescent plasma (**Figure 5A**). The analysis identified the same cut point when restricted to patients transfused with convalescent plasma with an anti-RBD IgG titer ≥1:1350. Therefore, we performed the propensity score-matched analysis using this cut point as a restrictor. Cohorts were again balanced across all matching criteria (data not shown). The resulting Kaplan-Meier curves showed significantly decreased mortality within 60 days post-Day 0 in the cohort transfused with convalescent plasma with an anti-RBD IgG ≥1:1350 within 44 hrs of admission relative to propensity score-matched controls (*P*=0.004) (**Figure 5B**). Point estimates of the outcomes for the analysis restricted to transfusion of high titer convalescent plasma within 44 hrs confirm these findings (**Table 3**). Univariate Cox regression in this cohort revealed a significant unadjusted HR for mortality within 60 days (HR=3.26 for controls, *P*=0.01). Likewise, multivariate Cox regression showed a significant adjusted HR for mortality within 28 days (aHR=2.63 for controls, *P*=0.04) and within 60 days post-Day 0 (aHR = 2.90 for controls, *P*=0.02) (**Table 4**).

**Figure 5.**
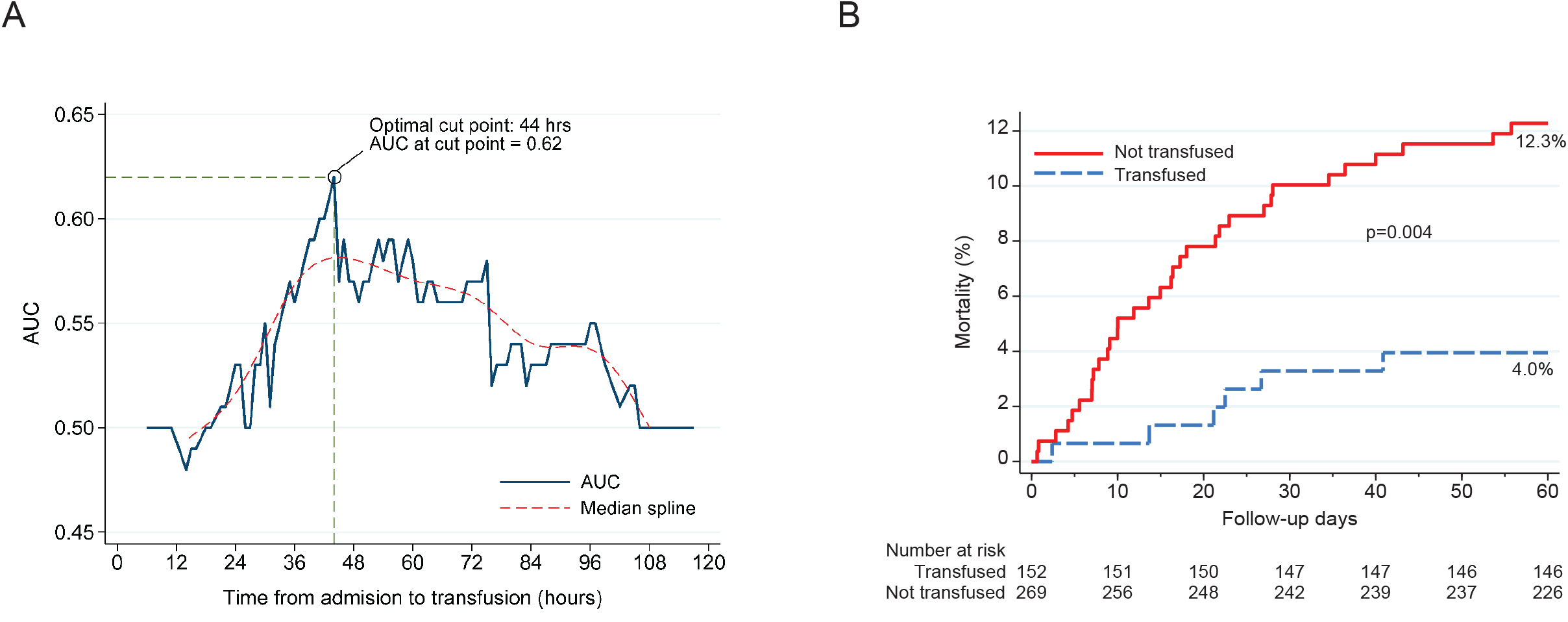
A) ROC curve with Youden index analysis for mortality within 60 days shown for all patients transfused with COVID-19 convalescent plasma. Optimal cut point identified as 44 hours with an area under the curve (AUC) of 0.62. Youden index was 0.23 with standard error of 0.0926. Sensitivity at cut point was 0.75 with a specificity of 0.48. B) Kaplan-Meier curves for mortality within 60 days post-Day 0 for patients transfused with plasma with an anti-RBD IgG titer ≥1:1350 within 44 hours after admission (blue) propensity score-matched to control patients (red).

**Table 3.** Outcome summary, Propensity score-matched, Transfused with Plasma with Titer ≥1:1350 within 44 hours of Admission

| | Propensity score-matched, transfused $\leq 44$ hrs, Titer $\geq 1350$ | | | | |
| --- | --- | --- | --- | --- | --- |
|  | Total<br>(N=421) | Not Transfused<br>(n=269) | Transfused<br>(n=152) | Point estimate*<br>(95% CI) | P-value |
| Disposition, 60 days |  |  |  |  |  |
| Still admitted | 4 (1.0) | 1 (0.4) | 3 (2.0) | 0.20 (0.02, 2.00) | 0.17 |
| Discharge | 377 (89.5) | 234 (87.0) | 143 (94.1) | (base outcome) |  |
| Death | 40 (9.5) | 34 (12.6) | 6 (3.9) | 3.46 (1.40, 8.56) | **0.01** |
| Overall mortality with no time constraints |  |  |  |  |  |
| Alive | 381 (90.5) | 235 (87.4) | 146 (96.1) | 3.20 (1.36, 7.54) | **0.01** |
| Deceased | 40 (9.5) | 34 (12.6) | 6 (3.9) |  |  |
| Overall mortality within 28 days post-Day 0 |  |  |  |  |  |
| Alive | 390 (92.6) | 243 (90.3) | 147 (96.7) | 2.94 (1.12, 7.74) | **0.03** |
| Deceased | 31 (7.4) | 26 (9.7) | 5 (3.3) |  |  |
| Overall mortality within 60 days post-Day 0 |  |  |  |  |  |
| Alive | 382 (90.7) | 236 (87.7) | 146 (96.1) | 3.11 (1.29, 7.50) | **0.01** |
| Deceased | 39 (9.3) | 33 (12.3) | 6 (3.9) |  |  |
| Length of Stay post-Day 0, median (IQR) | 5.3 (2.9, 10.0) | 5.0 (2.7, 10.0) | 5.7 (3.5, 9.7) | -0.57 (-2.98, 1.85) | 0.65 |
| Required ICU post-Day 0 |  |  |  |  |  |
| No | 291 (69.1) | 183 (68.0) | 108 (71.1) | 1.10 (0.81, 1.50) | 0.52 |
| Yes | 130 (30.9) | 86 (32.0) | 44 (28.9) |  |  |
| ICU length of stay post-Day 0, mean (±SD) | 10.8 (±11.3) | 10.7 (±11.3) | 11.2 (±11.4) | -0.52 (-4.55, 3.50) | 0.80 |
| Mechanical ventilation requirement, post-Day 0 |  |  |  |  |  |
| No | 347 (82.4) | 217 (80.7) | 130 (85.5) | 1.34 (0.86, 2.08) | 0.20 |
| Yes | 74 (17.6) | 52 (19.3) | 22 (14.5) |  |  |
| Mechanical ventilation days post-Day 0 (for patients that required ventilation), mean (±SD) | 19.8 (±19.2) | 17.9 (±15.7) | 24.2 (±25.5) | -6.34 (-16.72, 4.03) | 0.23 |
| Supplemental oxygen post-Day 0 |  |  |  |  |  |
| No | 60 (14.3) | 50 (18.6) | 10 (6.6) | 0.79 (0.75, 0.84) | **<0.001** |
| Yes | 361 (85.7) | 219 (81.4) | 142 (93.4) |  |  |
| Supplemental oxygen days post-Day 0 (for patients that required sup. oxygen), median (IQR) | 5.4 (±5.8) | 5.6 (±6.4) | 5.0 (±4.8) | 0.60 (-0.55, 1.76) | 0.31 |
| Ventilation status at Day 7 |  |  |  |  |  |
| Room air | 272 (64.6) | 172 (63.9) | 100 (65.8) | (base outcome) |  |
| Low flow | 38 (9.0) | 19 (7.1) | 19 (12.5) | 0.58 (0.29, 1.15) | 0.12 |
| High flow/NIPPV | 47 (11.2) | 34 (12.6) | 13 (8.6) | 1.52 (0.77, 3.01) | 0.23 |
| Mechanical ventilation | 52 (12.4) | 35 (13.0) | 17 (11.2) | 1.20 (0.64, 2.25) | 0.58 |
| ECMO | 4 (1.0) | 2 (0.7) | 2 (1.3) | 0.58 (0.08, 4.22) | 0.59 |
| Death | 8 (1.9) | 7 (2.6) | 1 (0.7) | 4.07 (0.49, 33.99) | 0.20 |
| Ventilation status at Day 14 |  |  |  |  |  |
| Room air | 346 (82.2) | 214 (79.6) | 132 (86.8) | (base outcome) |  |
| Low flow | 4 (1.0) | 2 (0.7) | 2 (1.3) | 0.62 (0.09, 4.43) | 0.63 |
| High flow/NIPPV | 13 (3.1) | 10 (3.7) | 3 (2.0) | 2.06 (0.55, 7.69) | 0.28 |
| Mechanical ventilation | 37 (8.8) | 25 (9.3) | 12 (7.9) | 1.29 (0.62, 2.65) | 0.50 |
| ECMO | 3 (0.7) | 2 (0.7) | 1 (0.7) | 1.23 (0.11, 13.79) | 0.87 |
| Death | 18 (4.3) | 16 (5.9) | 2 (1.3) | 4.93 (1.09, 22.38) | **0.04** |
| Ventilation status at Day 28 |  |  |  |  |  |
| Room air | 363 (86.2) | 223 (82.9) | 140 (92.1) | -- | -- |
| Low flow | 4 (1.0) | 4 (1.5) | 0 (0.0) | -- | -- |
| High flow/NIPPV | 3 (0.7) | 3 (1.1) | 0 (0.0) | -- | -- |
| Mechanical ventilation | 18 (4.3) | 12 (4.5) | 6 (3.9) | -- | -- |
| ECMO | 2 (0.5) | 1 (0.4) | 1 (0.7) | -- | -- |
| Death | 31 (7.4) | 26 (9.7) | 5 (3.3) | -- | -- |
| Ventilation status at Day 60 |  |  |  |  |  |
| Room air | 376 (89.3) | 233 (86.6) | 143 (94.1) | (base outcome) |  |
| Low flow |  | 0 (0.0) | 0 (0.0) | -- | -- |
| High flow/NIPPV |  | 0 (0.0) | 0 (0.0) | -- | -- |
| Mechanical ventilation | 6 (1.4) | 3 (1.1) | 3 (2.0) | 0.61 (0.12, 3.10) | 0.55 |
| ECMO |  | 0 (0.0) | 0 (0.0) | -- | -- |
| Death | 39 (9.3) | 33 (12.3) | 6 (3.9) | 3.38 (1.33, 8.55) | **0.01** |
| Clinical Improvement relative to Day 0 at Day 7 |  |  |  |  |  |
| No | 142 (33.7) | 97 (36.1) | 45 (29.6) | 0.91 (0.79, 1.05) | 0.19 |
| Yes | 279 (66.3) | 172 (63.9) | 107 (70.4) |  |  |
| Clinical Improvement relative to Day 0 at Day 14 |  |  |  |  |  |
| No | 81 (19.2) | 62 (23.0) | 19 (12.5) | 0.76 (0.71, 0.82) | **<0.001** |
| Yes | 340 (80.8) | 207 (77.0) | 133 (87.5) |  |  |
| Clinical Improvement relative to Day 0 at Day 28 |  |  |  |  |  |
| No | 65 (15.4) | 53 (19.7) | 12 (7.9) | 0.87 (0.81, 0.94) | **0.001** |
| Yes | 356 (84.6) | 216 (80.3) | 140 (92.1) |  |  |
| Clinical Improvement relative to Day 0 at Day 60 |  |  |  |  |  |
| No | 54 (12.8) | 45 (16.7) | 9 (5.9) | 0.89 (0.83, 0.95) | **<0.001** |
| Yes | 367 (87.2) | 224 (83.3) | 143 (94.1) |  |  |
| Interleukin-6 delta (Day 7-Day 0), (pg/mL), median (IQR) | 36.0 (-52.0, 370.0) | 16.5 (-39.0, 336.0) | 39.0 (-56.5, 531.0) | -151.40 (-591.29, 288.49) | 0.50 |
| C-reactive protein delta (Day 7-Day 0), (mg/dL), median (IQR) | -10.8 (-19.7, -5.0) | -10.8 (-22.3, -4.9) | -10.7 (-19.3, -5.3) | -0.09 (-3.90, 3.71) | 0.96 |
| Ferritin delta (Day 7-Day 0), (ng/mL), median (IQR) | -75.0 (-419.0, 192.0) | -110.0 (-468.0, 205.0) | -51.0 (-323.0, 189.0) | -29.53 (-376.08, 317.02) | 0.87 |
| Fibrinogen delta (Day 7-Day 0), (mg/dL), median (IQR) | -239.0 (-361.0, -74.5) | -140.0 (-321.5, -73.5) | -302.5 (-378.0, -82.5) | 55.24 (-41.82, 152.29) | 0.27 |
| D-dimer delta (Day 7-Day 0), (µg/mL FEU), median (IQR) | 0.4 (-0.3, 2.7) | 0.4 (-0.2, 3.4) | 0.3 (-0.3, 1.7) | 1.13 (-0.50, 2.76) | 0.18 |

Values are in median (interquartile range, IQR) for continuous variables and number and % for categorical variables
\*Point estimate obtained from the generalized linear models (GLM, for binary and continuous dependent variables) or multinomial logistic regression (for categorical variables) which is risk ratio of outcome in non-transfusion versus transfusion (if categorical outcomes or coefficient of outcome in non-transfusion versus transfusion (if continuous outcomes)
CI: Confidence Interval ECMO: extracorporeal membrane oxygenation; FEU: fibrinogen equivalent units; NIPPV: noninvasive positive pressure ventilation

**Table 4.** Univariate and Multivariate Cox Regression, Overall Mortality within 28 and 60 days, Controls Matched to Cases that Received Plasma with Titer ≥1:1350 within 44 hours of Hospital Admission

| Univariate | Within 60 days |  |  |  |
| --- | --- | --- | --- | --- |
|  | Alive<br>(n=382) | Deceased<br>(n=39) | Unadjusted HR<br>(95% CI) | P-value |
| Convalescent plasma transfusion |  |  |  |  |
| Transfused | 146 (38.2) | 6 (15.4) | (reference) |  |
| Not Transfused | 236 (61.8) | 33 (84.6) | 3.26 (1.32, 8.04) | **0.01** |
| Age (years), median (IQR) | 51.0 (39.0, 60.0) | 65.0 (59.0, 75.0) | 1.08 (1.06, 1.11) | **<0.001** |
| Age (years) |  |  |  |  |
| <30 | 28 (7.3) | 1 (2.6) | 1.47 (0.13, 16.32) | 0.75 |
| 30-39 | 68 (17.8) | 0 (0.0) | -- | -- |
| 40-49 | 83 (21.7) | 2 (5.1) | (reference) |  |
| 50-59 | 104 (27.2) | 7 (17.9) | 2.71 (0.56, 13.24) | 0.22 |
| 60-69 | 73 (19.1) | 15 (38.5) | 7.80 (1.74, 34.96) | **0.01** |
| 70-79 | 20 (5.2) | 9 (23.1) | 16.08 (3.38, 76.64) | **<0.001** |
| $\geq 80$ | 6 (1.6) | 5 (12.8) | 26.69 (5.03, 141.70) | **<0.001** |
| Sex |  |  |  |  |
| Female | 160 (41.9) | 11 (28.2) | (reference) |  |
| Male | 222 (58.1) | 28 (71.8) | 1.76 (0.86, 3.59) | 0.12 |
| Race |  |  |  |  |
| White | 250 (65.4) | 30 (76.9) | (reference) |  |
| Black | 78 (20.4) | 5 (12.8) | 0.55 (0.21, 1.42) | 0.21 |
| Asian | 27 (7.1) | 2 (5.1) | 0.63 (0.15, 2.77) | 0.54 |
| Other | 13 (3.4) | 0 (0.0) | -- | -- |
| Unknown | 14 (3.7) | 2 (5.1) | 1.18 (0.27, 5.09) | 0.82 |
| Ethnicity |  |  |  |  |
| Non-Hispanic | 176 (46.1) | 20 (51.3) | (reference) |  |
| Hispanic | 201 (52.6) | 19 (48.7) | 0.83 (0.45, 1.53) | 0.56 |
| Unknown | 5 (1.3) | 0 (0.0) |  |  |
| Body mass index (kg/m<sup>2</sup>), median (IQR) | 31.6 (28.3, 36.8) | 30.2 (25.9, 33.6) | 0.94 (0.89, 0.99) | **0.02** |
| Body mass index (kg/m<sup>2</sup>) |  |  |  |  |
| <18.5 | 1 (0.3) | 0 (0.0) | -- | -- |
| 18.5-24.9 | 30 (7.9) | 7 (17.9) | (reference) |  |
| 25-29.9 | 114 (29.8) | 12 (30.8) | 0.47 (0.18, 1.24) | 0.13 |
| ≥30 | 237 (62.0) | 20 (51.3) | 0.38 (0.17, 0.85) | **0.02** |
| Body mass index ≥30 (kg/m<sup>2</sup>) |  |  |  |  |
| <30 | 145 (38.0) | 19 (48.7) | (reference) |  |
| ≥30 | 237 (62.0) | 20 (51.3) | 0.65 (0.36, 1.19) | 0.17 |
| Hypertension |  |  |  |  |
| No | 199 (52.1) | 10 (25.6) | (reference) |  |
| Yes | 183 (47.9) | 29 (74.4) | 2.98 (1.41, 6.30) | **0.004** |
| Diabetes |  |  |  |  |
| No | 237 (62.0) | 16 (41.0) | (reference) |  |
| Yes | 145 (38.0) | 23 (59.0) | 2.23 (1.26, 3.97) | **0.01** |
| Chronic pulmonary disease |  |  |  |  |
| No | 344 (90.1) | 32 (82.1) | (reference) |  |
| Yes | 38 (9.9) | 7 (17.9) | 1.88 (0.84, 4.23) | 0.13 |
| Chronic kidney disease |  |  |  |  |
| No | 357 (93.5) | 34 (87.2) | (reference) |  |
| Yes | 25 (6.5) | 5 (12.8) | 1.93 (0.80, 4.67) | 0.15 |
| Hyperlipidemia |  |  |  |  |
| No | 281 (73.6) | 21 (53.8) | (reference) |  |
| Yes | 101 (26.4) | 18 (46.2) | 2.26 (1.19, 4.29) | **0.01** |
| Coronary disease |  |  |  |  |
| No | 367 (96.1) | 36 (92.3) | (reference) |  |
| Yes | 15 (3.9) | 3 (7.7) | 2.00 (0.60, 6.59) | 0.26 |
| Baseline outcome group |  |  |  |  |
| Room air | 12 (3.1) | 1 (2.6) | (reference) |  |
| Supplemental oxygen | 343 (89.8) | 24 (61.5) | 0.83 (0.10, 6.52) | 0.86 |
| Mechanical ventilation | 27 (7.1) | 14 (35.9) | 5.03 (0.69, 36.51) | 0.11 |
| Ventilation status at Day 0 |  |  |  |  |
| Room air | 33 (8.6) | 1 (2.6) | (reference) |  |
| Low flow | 248 (64.9) | 8 (20.5) | 1.05 (0.14, 8.10) | 0.97 |
| High flow/NIPPV | 72 (18.8) | 17 (43.6) | 7.01 (0.96, 51.32) | 0.06 |
| Mechanical ventilation | 27 (7.1) | 13 (33.3) | 12.98 (1.92, 87.59) | **0.01** |
| ECMO | 2 (0.5) | 0 (0.0) | -- | -- |
| ABO blood group |  |  |  |  |
| A | 82 (26.1) | 11 (29.7) | 1.16 (0.58, 2.34) | 0.68 |
| B | 43 (13.7) | 4 (10.8) | 0.83 (0.28, 2.48) | 0.74 |
| AB | 9 (2.9) | 1 (2.7) | 0.95 (0.13, 6.95) | 0.96 |
| O | 180 (57.3) | 21 (56.8) | (reference) |  |
| Rh blood group |  |  |  |  |
| Negative | 31 (9.9) | 7 (18.9) | (reference) |  |
| Positive | 283 (90.1) | 30 (81.1) | 0.50 (0.24, 1.08) | 0.08 |
| Interleukin-6 at baseline, (pg/mL), median (IQR) (**n=316**) | 57.0 (25.0, 116.5) | 85.5 (52.0, 192.5) | 1.001 (1.00, 1.001) | **<0.001** |
| Interleukin-6 delta (Day 7-baseline), (pg/mL), median (IQR) (**n=98**) | 3.5 (-52.0, 296.0) | 323.5 (-12.5, 1101.5) | 1.00 (1.00, 1.00) | 0.65 |
| C-reactive protein at baseline, (mg/dL), median (IQR) (**n=353**) | 9.7 (5.6, 16.6) | 12.7 (5.5, 19.1) | 1.02 (0.98, 1.05) | 0.31 |
| C-reactive protein delta (Day 7-baseline), (mg/dL), median (IQR) (**n=169**) | -10.9 (-19.7, -5.4) | -7.0 (-20.2, -4.3) | 1.00 (0.97, 1.03) | 0.92 |
| Ferritin at baseline, (ng/mL), median (IQR) (**n=358**) | 791.0 (375.0, 1462.0) | 1408.0 (509.0, 2152.0) | 1.0001 (1.00, 1.0001) | **<0.001** |
| Ferritin delta (Day 7-baseline), (ng/mL), median (IQR) (**n=163**) | -77.0 (-438.0, 174.0) | -66.0 (-322.0, 314.0) | 1.00 (1.00, 1.00) | 0.11 |
| Fibrinogen at baseline, (mg/dL), median (IQR) (**n=287**) | 643.0 (535.0, 748.0) | 637.0 (589.0, 712.0) | 1.00 (0.99, 1.00) | 0.23 |
| Fibrinogen delta (Day 7-baseline), (mg/dL), median (IQR) (**n=60**) | -191.0 (-360.0, -69.0) | -248.0 (-477.0, -141.0) | 1.00 (1.00, 1.00) | 0.22 |
| D-dimer at baseline, (µg/mL FEU), median (IQR) (**n=364**) | 0.9 (0.6, 1.7) | 2.0 (0.8, 4.3) | 1.15 (1.10, 1.21) | **<0.001** |
| D-dimer delta (Day 7-baseline), (µg/mL FEU), median (IQR) (**n=174**) | 0.2 (-0.3, 1.6) | 3.2 (0.7, 13.6) | 1.08 (1.04, 1.14) | **0.001** |
| Concomitant medication |  |  |  |  |
| Any steroids | 232 (60.7) | 37 (94.9) | 11.03 (2.69, 45.28) | **0.001** |
| Dexamethasone | 129 (33.8) | 16 (41.0) | 1.34 (0.67, 2.67) | 0.41 |
| Hydrocortisone | 9 (2.4) | 16 (41.0) | 13.97 (7.70, 25.35) | **<0.001** |
| Methylprednisolone | 143 (37.4) | 28 (71.8) | 3.85 (1.84, 8.09) | **<0.001** |
| Prednisone | 26 (6.8) | 4 (10.3) | 1.48 (0.54, 4.06) | 0.44 |
| Azithromycin | 265 (69.4) | 30 (76.9) | 1.41 (0.69, 2.86) | 0.35 |
| Hydroxychloroquine | 35 (9.2) | 3 (7.7) | 0.83 (0.25, 2.74) | 0.76 |
| Lopinavir/ritonavir | 1 (0.3) | 0 (0.0) | -- | -- |
| Remdesivir | 155 (40.6) | 15 (38.5) | 0.91 (0.47, 1.79) | 0.79 |
| Ribavirin | 4 (1.0) | 0 (0.0) | -- | -- |
| Tocilizumab | 158 (41.4) | 31 (79.5) | 5.01 (2.35, 10.68) | **<0.001** |
| **Multivariate (n=421)** | **Within 28 days** |  | **Within 60 days** |  |
|  | **Adjusted HR**<br>**(95% CI)** | **P-value** | **Adjusted HR**<br>**(95% CI)** | **P-value** |
| Convalescent plasma transfusion |  |  |  |  |
| Transfused | (reference) |  | (reference) |  |
| Not Transfused | 2.63 (1.04, 6.64) | **0.04** | 2.90 (1.22, 6.94) | **0.02** |
| Age (years) | 1.09 (1.06, 1.13) | **<0.001** | 1.08 (1.05, 1.12) | **<0.001** |
| Diabetes | 1.74 (0.90, 3.38) | 0.10 | 1.87 (1.04, 3.36) | **0.04** |
| Any steroid | 8.45 (1.78, 40.03) | **0.01** | 11.16 (2.54, 48.99) | **0.001** |
| C-statistic | **C-statistic = 0.86** | -- | **C-statistic = 0.86** | -- |

Values are in median (interquartile range, IQR) for continuous variables and number (%) for categorical variables.
Steroids and tocilizumab were treated as time-varying covariates in the multivariate model.
CI: confidence interval; HR: hazard ratio; ECMO: extracorporeal membrane oxygenation; FEU: fibrinogen equivalent units; NIPPV: noninvasive positive pressure ventilation

## DISCUSSION

Transfusion of convalescent plasma has emerged in the last six months as a promising therapy for COVID-19 patients and has been granted emergency use authorization for hospitalized patients by the FDA. Because of the logistical challenges of planning and executing a study during a rapidly changing pandemic involving very complex medical patients, the results of few completed controlled studies assessing convalescent plasma efficacy have been published. Here, we provide an analysis of a propensity score-matched study from a large cohort of hospitalized COVID-19 patients who were transfused in one healthcare system with high-titer convalescent plasma qualified in one laboratory. In the aggregate, the data confirm and extend findings from our interim analysis suggesting that transfusion of convalescent plasma with high titer anti-RBD IgG is safe and significantly decreases COVID-19 mortality.^3^ Transfusion later in hospitalization or later in the disease course (e.g., post-intubation) had no significant benefit on mortality, regardless of plasma titer. Several lines of evidence support our findings, including survival analyses of specific cohorts of transfused patients relative to matched controls, point estimates from the generalized linear model and multinomial logistic regression, and univariate and multivariate analyses.

The current analysis addressed several limitations we identified in our interim analysis.^3^ First, the patient sample size is almost three times as large as that included in our interim analysis. Second, we included additional covariates in the propensity score matching algorithm, including relevant concomitant medications (any steroid, azithromycin, hydroxychloroquine, remdesivir, ribavirin, and tocilizumab). Importantly, factors identified as having a significant adjusted HR for mortality for all hospitalized COVID-19 patients were included in the propensity match. Third, because a large proportion of deaths occurred after 28 days post-Day 0, we assessed a 60-day outcome. Fourth, control patients enrolled in other clinical trials involving alternative experimental therapies were excluded. Fifth, when possible, we performed multivariate analyses assessing factors associated with mortality within 60 days. Finally, we used ROC analysis with Youden index to identify the optimal cut point at which transfusion of convalescent plasma is most useful with respect to altering mortality.

Our results bear on other recent studies treating patients with convalescent plasma.^4, 9, 13-16^ For example, a recent fixed-effect meta-analysis model assessing 12 controlled studies of COVID-19 convalescent plasma found that the aggregate mortality rate of transfused COVID-19 patients was significantly lower than that of non-transfused patients.^4^ Results from three randomized controlled studies and one large observational study have recently been released.^2, 17-19^ The PLACID trial found convalescent plasma was not associated with significantly reduced mortality or progression to severe disease.^17^ However, resolution of shortness of breath, fatigue, and negative conversion of SARS-CoV-2 viral RNA at Day 7 was higher in the transfused study arm. The authors acknowledged several limitations of their study. For example, the proportion of patients with comorbidities, especially diabetes, was higher in the transfused study arm. Importantly, most of the convalescent plasma donors were young with mild disease and their median titer of neutralizing antibody was 1:40, a value considerably lower than the FDA-recommended neutralizing antibody titer of 1:160. In addition, neutralizing antibody titers were not determined before transfusion, which means the highest titer units were not used for transfusion. Similar results were reported for a randomized controlled trial conducted in Chile in which neutralizing antibody titers in donor plasma were not determined prior to transfusion.^19^ In contrast, interim analysis of a randomized controlled trial from Spain with 81 randomized patients, reported that no patients progressed to mechanical ventilation or death among the 38 patients receiving convalescent plasma (0%), whereas six of 43 patients (14%) in the control arm did.^18^ Mortality rates were 0% versus 9.3% at Days 15 and 29 for the active and control groups, respectively. All transfused convalescent plasma units had neutralizing antibodies with a titer >1:80 with a median titer of 1:292. Unfortunately, the trial was stopped after the first interim analysis due to decreased recruitment related to better control of the pandemic. In contrast to several of the studies cited above, we methodically selected units for transfusion based on the ELISA data identifying the highest level of IgG antibody directed against spike ectodomain and RBD. We transfused compatible donor units determined to have the highest antibody titer available, an approach confirmed by our retrospective assessment of anti-SARS-CoV-2 IgG by the Ortho VITROS assay (**Figure 1**). Thus, the vast majority of our patients were transfused with convalescent plasma units with very high titer anti-spike protein IgG. We think it reasonable to speculate that this strategy contributed to differences in outcomes observed between our study and several others that did not transfuse patients with plasma units specifically chosen to have very high IgG antibody levels against spike protein. Overall, the results from various published studies highlight the difficulty in drawing definitive conclusions for convalescent plasma efficacy from multiple studies with variable design, a problem that can extend to and thereby hobble randomized controlled trials with different study designs.

Substantial efforts to collect, use, and study COVID-19 convalescent plasma continue worldwide. Our study has several implications for these efforts. The data presented here may inform the design and conduct of ongoing or future studies. For example, we conclude that transfusing plasma units with low or no antibody titer against spike protein is unlikely to be beneficial. Our data support the concept that assessment of antibody titer by either a viral neutralization assay or a surrogate thereof prior to transfusion is essential, regardless of the type of trial being conducted. In addition, transfusing relatively soon after hospitalization will be more beneficial than the alternative. Our finding that a large proportion of deaths in COVID-19 patients occurs after Day 28 may also have implications for study design, as findings at Day 28 may not apply over a longer follow-up period.

Importantly, our study has several limitations. First, it is a propensity score-matched study rather than a randomized controlled trial. Although we made every effort to control for all important covariates, potentially relevant covariates may have been omitted unintentionally from the matching algorithm. Second, the background standard of care for COVID-19 has evolved as new data emerged. Thus, we may not have completely addressed the potential for variations over time in background standard of care and period effect as sources of confounding in our dataset. Third, there was heterogeneity in the transfusion of two units versus one based on inventory limitations early in the study and on patient enrollment in other trials that specifically excluded redosing of convalescent plasma. Fourth, our analysis was based on patient data available in the electronic medical record. Fifth, we note that the results reflect the experience of one system of eight hospitals in the Houston metropolitan region that have a fairly uniform approach to COVID-19 patient care. Our findings may not apply to all hospitalized COVID-19 patients because of inter-institutional and/or regional heterogeneity in medical care. Sixth, baseline inflammatory marker measurements were not included in the matching algorithm due to the high proportion of missing data points. Our study approach facilitated rapid assessment of safety and efficacy of high-titer anti-SARS-CoV-2 convalescent plasma transfusion during early phases of a rapidly evolving pandemic with uncertain trajectory. The data presented here may help to inform the science and logistics of ongoing and future studies that address the use of convalescent plasma for other emerging and rapidly disseminating infectious diseases.

To summarize, this propensity score-matched analysis of a large patient cohort confirms and extends our previous findings and suggests that transfusion of convalescent plasma containing very high titer anti-RBD IgG early in hospitalization reduces mortality in COVID-19 patients.

## Data Availability

Data available within the article or its supplementary materials.

## ACKNOWLEDGMENTS

We are deeply indebted to all of our volunteer plasma donors for their time, their generous gift, and their solidarity. We thank Katharine G. Dlouhy, Curt Hampton, and their team of coordinators and recruiters for outstanding efforts; Monisha Dey, Cheryl Chavez-East, who were instrumental in efficiently managing the donor center; Kate Cody, Sayali Kelkar, Belimat Askary, and the clinical analytics team for their assistance with data acquisition and management; Drs. Jessica Thomas and Zejuan Li, Erika Walker, the very talented and dedicated molecular technologists, and the many labor pool volunteers in the Molecular Diagnostics Laboratory for their dedication to patient care; the many donor center and blood bank phlebotomists and technologists for their dedication to donor and blood safety; Sasha Pejerrey, Adrienne Winston, and Heather McConnell for editorial assistance; Claude Moussa, Heather Patton, and other members of our laboratory information technology team for rapidly implementing the necessary electronic workflows; Pamela McShane, Dilzi Mody, and members of the biorepository team for their meticulous management of patient samples; Christina Talley, Susan Miller, and Mary Clancy for consistent, thorough, and outstanding advice; and Zivko Nikolov, Susan Woodard, and Michael Johanson at the National Center for Therapeutics Manufacturing at Texas A&M University for production of spike protein antigens. We express our gratitude to Manuel Hinojosa and Mark Vassallo for their extensive efforts to rapidly procure resources. We are indebted to Drs. Marc Boom and Dirk Sostman for their support, and to many very generous Houston citizens and businesses for their tremendous philanthropic support of this ongoing project, including but not limited to anonymous, Ann and John Bookout III, Carolyn and John Bookout, Ting Tsung and Wei Fong Chao Foundation, Ann and Leslie Doggett, Freeport LNG, the Hearst Foundations, Jerold B. Katz Foundation, C. James and Carole Walter Looke, Diane and David Modesett, the Sherman Foundation, Paula and Joseph C. “Rusty” Walter III, and Aramco Americas. Dr. Jason S. McLellan (University of Texas at Austin) provided the mAb CR3022 and the spike protein expression vectors, and we thank the members of the Center for Systems and Synthetic Biology at the University of Texas at Austin for technical assistance. We thank Terumo BCT for continuously and rapidly supplying blood collection devices and supplies. We also thank Shmuel Shoham, MD for graciously sharing a draft study protocol for adaptation early in the study planning phase.

Statement of ethical assurance: JMM is the guarantor of this work and, as such, had full access to all of the data in the study and takes responsibility for the integrity of the data and the accuracy of the data analysis.

## Notes

### Competing Interest Statement

ES is the local principal investigator for a clinical trial sponsored by Regeneron assessing an investigational therapy for COVID-19.

### Funding Statement

This study was supported by the Fondren Foundation, Houston Methodist Hospital and Research Institute (to JMM).

### Author Declarations

Houston Methodist Research Institute ethics review board

