## Supplemental Table 1 for "Early transfusion of a large cohort of COVID-19 patients with high titer anti-SARS-CoV-2 spike protein IgG convalescent plasma confirms a signal of significantly decreased mortality"

**Table S1. Patient Characteristics in All Patients who Met 60-day Outcome**

|  | **Total** | **Not Transfused** | **Transfused** | **P-value** |
| --- | --- | --- | --- | --- |
|  | **(N=3468)** | **(*n*=3117)** | **(*n*=351)** |  |
| Age (years), median (IQR) | 59.0 (47.0, 71.0) | 60.0 (48.0, 72.0) | 53.0 (43.0, 62.0) | **<0.001** |
| Age (years) |  |  |  | **<0.001** |
| <30 | 211 (6.1) | 192 (6.2) | 19 (5.4) |  |
| 30-39 | 337 (9.7) | 284 (9.1) | 53 (15.1) |  |
| 40-49 | 493 (14.2) | 419 (13.4) | 74 (21.1) |  |
| 50-59 | 727 (21.0) | 636 (20.4) | 91 (25.9) |  |
| 60-69 | 711 (20.5) | 635 (20.4) | 76 (21.7) |  |
| 70-79 | 592 (17.1) | 561 (18.0) | 31 (8.8) |  |
| ≥80 | 397 (11.4) | 390 (12.5) | 7 (2.0) |  |
| Sex |  |  |  | **0.01** |
| Female | 1708 (49.3) | 1557 (50.0) | 151 (43.0) |  |
| Male | 1760 (50.7) | 1560 (50.0) | 200 (57.0) |  |
| Race |  |  |  | **<0.001** |
| White | 2159 (62.3) | 1933 (62.0) | 226 (64.4) |  |
| Black | 947 (27.3) | 877 (28.1) | 70 (19.9) |  |
| Asian | 153 (4.4) | 130 (4.2) | 23 (6.6) |  |
| Other | 90 (2.6) | 75 (2.4) | 15 (4.3) |  |
| Unknown | 119 (3.4) | 102 (3.3) | 17 (4.8) |  |
| Ethnicity |  |  |  | **<0.001** |
| Non-Hispanic | 2098 (60.5) | 1933 (62.0) | 165 (47.0) |  |
| Hispanic | 1325 (38.2) | 1142 (36.6) | 183 (52.1) |  |
| Unknown | 45 (1.3) | 42 (1.3) | 3 (0.9) |  |
| Body mass index (kg/m^2^), median (IQR) | 30.1 (25.9, 35.6) | 29.9 (25.7, 35.4) | 31.7 (27.5, 36.9) | **<0.001** |
| Body mass index (kg/m^2^) |  |  |  | **<0.001** |
| <18.5 | 41 (1.2) | 40 (1.3) | 1 (0.3) |  |
| 18.5-24.9 | 643 (18.9) | 607 (19.9) | 36 (10.3) |  |
| 25-29.9 | 996 (29.3) | 894 (29.3) | 102 (29.1) |  |
| ≥30 | 1725 (50.7) | 1513 (49.5) | 212 (60.4) |  |
| Body mass index ≥30 (kg/m^2^) |  |  |  | **<0.001** |
| <30 | 1680 (49.3) | 1541 (50.5) | 139 (39.6) |  |
| ≥30 | 1725 (50.7) | 1513 (49.5) | 212 (60.4) |  |
| Hypertension |  |  |  | 0.11 |
| No | 1617 (46.6) | 1439 (46.2) | 178 (50.7) |  |
| Yes | 1851 (53.4) | 1678 (53.8) | 173 (49.3) |  |
| Diabetes |  |  |  | 0.11 |
| No | 1956 (56.4) | 1744 (56.0) | 212 (60.4) |  |
| Yes | 1512 (43.6) | 1373 (44.0) | 139 (39.6) |  |
| Chronic pulmonary disease |  |  |  | **0.01** |
| No | 2912 (84.0) | 2600 (83.4) | 312 (88.9) |  |
| Yes | 556 (16.0) | 517 (16.6) | 39 (11.1) |  |
| Chronic kidney disease |  |  |  | **<0.001** |
| No | 2678 (77.2) | 2367 (75.9) | 311 (88.6) |  |
| Yes | 790 (22.8) | 750 (24.1) | 40 (11.4) |  |
| Hyperlipidemia |  |  |  | **0.01** |
| No | 2171 (62.6) | 1928 (61.9) | 243 (69.2) |  |
| Yes | 1297 (37.4) | 1189 (38.1) | 108 (30.8) |  |
| Coronary disease |  |  |  | **<0.001** |
| No | 2922 (84.3) | 2596 (83.3) | 326 (92.9) |  |
| Yes | 546 (15.7) | 521 (16.7) | 25 (7.1) |  |
| Baseline ventilation status (within 48 hours of admission) |  |  |  | **<0.001** |
| Room air | 881 (25.4) | 855 (27.4) | 26 (7.4) |  |
| Supplemental oxygen | 2382 (68.7) | 2078 (66.7) | 304 (86.6) |  |
| Mechanical ventilation | 205 (5.9) | 184 (5.9) | 21 (6.0) |  |
| ABO blood group |  |  |  | 0.32 |
| A | 768 (30.8) | 654 (30.5) | 114 (32.5) |  |
| B | 359 (14.4) | 316 (14.7) | 43 (12.3) |  |
| AB | 86 (3.4) | 78 (3.6) | 8 (2.3) |  |
| O | 1282 (51.4) | 1096 (51.1) | 186 (53.0) |  |
| Rh blood group |  |  |  | 0.35 |
| Negative | 223 (8.9) | 187 (8.7) | 36 (10.3) |  |
| Positive | 2272 (91.1) | 1957 (91.3) | 315 (89.7) |  |
| Interleukin-6 (pg/mL) at baseline, median (IQR) | 39.0 (16.0, 85.0) | 36.0 (15.0, 80.5) | 56.0 (28.5, 107.5) | **<0.001** |
| C-reactive protein (mg/dL) at baseline, median (IQR) | 8.0 (3.8, 14.6) | 7.8 (3.6, 14.5) | 9.3 (4.9, 15.4) | **0.02** |
| Ferritin (ng/mL) at baseline, median (IQR) | 621.0 (302.0, 1329.0) | 609.5 (292.0, 1327.0) | 713.0 (391.0, 1331.0) | 0.06 |
| Fibrinogen (mg/dL) at baseline, median (IQR) | 600.0 (497.0, 713.0) | 599.0 (494.0, 714.0) | 609.5 (520.0, 711.0) | 0.23 |
| D-dimer (µg/mL FEU) at baseline, median (IQR) | 0.9 (0.6, 1.8) | 1.0 (0.6, 1.9) | 0.8 (0.5, 1.3) | **<0.001** |
| Concomitant medication |  |  |  |  |
| Any steroids | 2127 (61.3) | 1882 (60.4) | 245 (69.8) | **<0.001** |
| Dexamethasone | 1558 (44.9) | 1410 (45.2) | 148 (42.2) | 0.27 |
| Hydrocortisone | 186 (5.4) | 162 (5.2) | 24 (6.8) | 0.20 |
| Methylprednisolone | 809 (23.3) | 665 (21.3) | 144 (41.0) | **<0.001** |
| Prednisone | 336 (9.7) | 286 (9.2) | 50 (14.2) | **0.002** |
| Azithromycin | 2095 (60.4) | 1838 (59.0) | 257 (73.2) | **<0.001** |
| Hydroxychloroquine | 415 (12.0) | 366 (11.7) | 49 (14.0) | 0.22 |
| Lopinavir/ritonavir | 40 (1.2) | 36 (1.2) | 4 (1.1) | 1.00 |
| Remdesivir | 622 (17.9) | 498 (16.0) | 124 (35.3) | **<0.001** |
| Ribavirin | 142 (4.1) | 122 (3.9) | 20 (5.7) | 0.11 |
| Tocilizumab | 714 (20.6) | 550 (17.6) | 164 (46.7) | **<0.001** |

Values are in median (interquartile range, IQR) for continuous variables and number (%) for categorical variables.

Differences between groups were compared using the Wilcoxon rank-sum test for continuous variables and Chi-square or Fisher’s exact tests for categorical variables, as appropriate.

FEU; fibrinogen equivalent units
