## Supplemental Table 2 for "Early transfusion of a large cohort of COVID-19 patients with high titer anti-SARS-CoV-2 spike protein IgG convalescent plasma confirms a signal of significantly decreased mortality"

**Table S2.** **Univariable Cox Regression, Overall Mortality Within 60 Days for All Patients Admitted**

|  | **Alive** | **Deceased** | **Unadjusted HR** | **P-value** |
| --- | --- | --- | --- | --- |
|  | **(*n*=4798)** | **(*n*=497)** | **(95% CI)** |  |
| Age (years), median (IQR) | 58.0 (45.0, 70.0) | 73.0 (63.0, 82.0) | 1.05 (1.05, 1.06) | **<0.001** |
| Age (years) |  |  |  |  |
| <30 | 353 (7.4) | 5 (1.0) | 0.64 (0.23, 1.75) | 0.39 |
| 30-39 | 495 (10.3) | 7 (1.4) | 0.64 (0.26, 1.55) | 0.32 |
| 40-49 | 708 (14.8) | 16 (3.2) | (reference) |  |
| 50-59 | 1020 (21.3) | 61 (12.3) | 2.58 (1.49, 4.48) | **0.001** |
| 60-69 | 974 (20.3) | 105 (21.1) | 4.55 (2.69, 7.69) | **<0.001** |
| 70-79 | 748 (15.6) | 151 (30.4) | 8.24 (4.92, 13.80) | **<0.001** |
| ≥80 | 500 (10.4) | 152 (30.6) | 12.40 (7.40, 20.75) | **<0.001** |
| Sex |  |  |  |  |
| Female | 2458 (51.2) | 210 (42.3) | (reference) |  |
| Male | 2340 (48.8) | 287 (57.7) | 1.39 (1.17, 1.66) | **<0.001** |
| Race |  |  |  |  |
| White | 3053 (63.6) | 312 (62.8) | (reference) |  |
| Black | 1235 (25.7) | 128 (25.8) | 0.99 (0.81, 1.21) | 0.92 |
| Asian | 225 (4.7) | 29 (5.8) | 1.23 (0.84, 1.80) | 0.28 |
| Other | 116 (2.4) | 15 (3.0) | 1.21 (0.72, 2.04) | 0.46 |
| Unknown | 169 (3.5) | 13 (2.6) | 0.76 (0.44, 1.33) | 0.34 |
| Ethnicity |  |  |  |  |
| Non-Hispanic | 2894 (60.3) | 344 (69.2) | (reference) |  |
| Hispanic | 1848 (38.5) | 147 (29.6) | 0.68 (0.56, 0.82) | **<0.001** |
| Unknown | 56 (1.2) | 6 (1.2) | 0.88 (0.39, 1.97) | 0.76 |
| Body mass index (kg/m^2^), median (IQR) | 30.2 (25.8, 35.7) | 28.5 (24.5, 34.0) | 0.97 (0.96, 0.99) | **<0.001** |
| Body mass index (kg/m^2^) |  |  |  |  |
| <18.5 | 65 (1.4) | 13 (2.6) | 1.43 (0.81, 2.53) | 0.22 |
| 18.5-24.9 | 890 (18.9) | 124 (25.1) | (reference) |  |
| 25-29.9 | 1344 (28.5) | 141 (28.5) | 0.76 (0.60, 0.97) | **0.03** |
| ≥30 | 2409 (51.2) | 217 (43.8) | 0.65 (0.52, 0.82) | **<0.001** |
| Body mass index ≥30 (kg/m^2^) |  |  |  |  |
| <30 | 2299 (48.8) | 278 (56.2) | (reference) |  |
| ≥30 | 2409 (51.2) | 217 (43.8) | 0.75 (0.63, 0.90) | **0.002** |
| Body mass index ≥35 (kg/m^2^) |  |  |  |  |
| <35 | 3423 (72.7) | 395 (79.8) | (reference) |  |
| ≥35 | 1285 (27.3) | 100 (20.2) | 0.68 (0.55, 0.85) | **0.001** |
| Body mass index ≥40 (kg/m^2^) |  |  |  |  |
| <40 | 4069 (86.4) | 451 (91.1) | (reference) |  |
| ≥40 | 639 (13.6) | 44 (8.9) | 0.62 (0.46, 0.85) | **0.003** |
| Hypertension |  |  |  |  |
| No | 2386 (49.7) | 196 (39.4) | (reference) |  |
| Yes | 2412 (50.3) | 301 (60.6) | 1.39 (1.16, 1.66) | **<0.001** |
| Diabetes |  |  |  |  |
| No | 2850 (59.4) | 200 (40.2) | (reference) |  |
| Yes | 1948 (40.6) | 297 (59.8) | 1.98 (1.66, 2.37) | **<0.001** |
| Chronic pulmonary disease |  |  |  |  |
| No | 4086 (85.2) | 378 (76.1) | (reference) |  |
| Yes | 712 (14.8) | 119 (23.9) | 1.70 (1.38, 2.08) | **<0.001** |
| Chronic kidney disease |  |  |  |  |
| No | 3863 (80.5) | 263 (52.9) | (reference) |  |
| Yes | 935 (19.5) | 234 (47.1) | 3.28 (2.75, 3.91) | **<0.001** |
| Hyperlipidemia |  |  |  |  |
| No | 3130 (65.2) | 231 (46.5) | (reference) |  |
| Yes | 1668 (34.8) | 266 (53.5) | 1.99 (1.67, 2.37) | **<0.001** |
| Coronary disease |  |  |  |  |
| No | 4140 (86.3) | 339 (68.2) | (reference) |  |
| Yes | 658 (13.7) | 158 (31.8) | 2.66 (2.20, 3.21) | **<0.001** |
| Baseline ventilation status (within 48 hours of admission) |  |  |  |  |
| Room air | 1425 (29.7) | 37 (7.4) | (reference) |  |
| Supplemental oxygen | 3212 (66.9) | 346 (69.6) | 3.86 (2.75, 5.42) | **<0.001** |
| Mechanical ventilation | 161 (3.4) | 114 (22.9) | 20.86 (14.39, 30.22) | **<0.001** |
| ABO blood group |  |  |  |  |
| A | 1005 (31.9) | 125 (29.8) | 0.91 (0.73, 1.14) | 0.43 |
| B | 455 (14.4) | 59 (14.1) | 0.95 (0.71, 1.26) | 0.71 |
| AB | 102 (3.2) | 16 (3.8) | 1.12 (0.67, 1.85) | 0.67 |
| O | 1592 (50.5) | 219 (52.3) | (reference) |  |
| Rh blood group |  |  |  |  |
| Negative | 270 (8.6) | 50 (11.9) | (reference) |  |
| Positive | 2884 (91.4) | 369 (88.1) | 0.71 (0.53, 0.95) | 0.02 |
| Interleukin-6 (pg/mL) at baseline, median (IQR) **(*n*=3360)** | 34.0 (14.0, 75.0) | 78.0 (36.0, 173.5) | 1.0002 (1.00, 1.0003) | **<0.001** |
| Interleukin-6 delta (Day 7-baseline), median (IQR**) (*n*=692)** | -5.0 (-54.0, 150.0) | 192.0 (-23.1, 891.0) | 1.0004 (1.00, 1.001) | **<0.001** |
| C-reactive protein (mg/dL) at baseline, median (IQR) **(*n*=3946)** | 7.6 (3.4, 14.1) | 12.2 (6.1, 19.2) | 1.03 (1.02, 1.04) | **<0.001** |
| C-reactive protein delta (Day 7-baseline), median (IQR) **(*n*=1345)** | -8.0 (-16.1, -2.6) | -5.7 (-14.2, 0.3) | 1.02 (1.01, 1.03) | **0.001** |
| Ferritin (ng/mL) at baseline, median (IQR) **(*n*=4031)** | 590.0 (286.0, 1230.0) | 842.0 (369.0, 1783.0) | 1.0001 (1.00, 1.0001) | **<0.001** |
| Ferritin delta (Day 7-baseline), median (IQR) **(*n*=1324)** | -26.0 (-298.0, 194.0) | 129.0 (-272.0, 562.0) | 1.002 (1.00, 1.0003) | **<0.001** |
| Fibrinogen (mg/dL) at baseline, median (IQR) **(*n*=3223)** | 601.0 (493.0, 709.0) | 584.0 (481.5, 712.0) | 0.99 (0.99, 1.00) | **0.02** |
| Fibrinogen delta (Day 7-baseline), median (IQR) **(*n*=496)** | -132.5 (-283.0, -8.0) | -86.0 (-249.0, 47.0) | 1.00 (1.00, 1.00) | 0.24 |
| D-dimer (µg/mL FEU) at baseline, median (IQR) **(*n*=4042)** | 0.9 (0.6, 1.8) | 1.7 (0.9, 3.9) | 1.11 (1.09, 1.13) | **<0.001** |
| D-dimer delta (Day 7-baseline), median (IQR) **(*n*=1312)** | -0.0 (-0.5, 0.7) | 1.1 (-0.3, 6.5) | 1.08 (1.05, 1.10) | <0.001 |
| Concomitant medication |  |  |  |  |
| Any steroids | 2901 (60.5) | 421 (84.7) | 3.38 (2.65, 4.32) | **<0.001** |
| Dexamethasone | 2321 (48.4) | 290 (58.4) | 1.47 (1.23, 1.75) | **<0.001** |
| Hydrocortisone | 101 (2.1) | 160 (32.2) | 11.38 (9.42, 13.76) | **<0.001** |
| Methylprednisolone | 893 (18.6) | 227 (45.7) | 3.22 (2.70, 3.85) | **<0.001** |
| Prednisone | 415 (8.6) | 70 (14.1) | 1.58 (1.23, 2.03) | <0.001 |
| Azithromycin | 2673 (55.7) | 293 (59.0) | 1.08 (0.90, 1.29) | 0.42 |
| Hydroxychloroquine | 367 (7.6) | 62 (12.5) | 1.56 (1.19, 2.03) | **0.001** |
| Lopinavir/ritonavir | 28 (0.6) | 14 (2.8) | 3.81 (2.24, 6.48) | **<0.001** |
| Remdesivir | 971 (20.2) | 109 (21.9) | 1.11 (0.90, 1.38) | 0.32 |
| Ribavirin | 104 (2.2) | 38 (7.6) | 3.09 (2.22, 4.31) | **<0.001** |
| Tocilizumab | 674 (14.0) | 193 (38.8) | 3.20 (2.67, 3.83) | **<0.001** |

Values are in median (interquartile range, IQR) for continuous variables and number (%) for categorical variables.

CI: confidence interval; FEU: fibrinogen equivalent units; HR: hazard ratio
