## Supplemental Table 3 for "Early transfusion of a large cohort of COVID-19 patients with high titer anti-SARS-CoV-2 spike protein IgG convalescent plasma confirms a signal of significantly decreased mortality"

| **Table S3. Multivariable Cox Regression, Overall Mortality within 60 Days for All Patients Admitted. (*N*=3538)** | | |
| --- | --- | --- |
|  | **Adjusted HR** | **P-value** |
|  | **(95% CI)** |  |
| Age (years) | 1.05 (1.04, 1.06) | **<0.001** |
| Male sex | 1.39 (1.14, 1.71) | **0.001** |
| Body mass index (kg/m^2^) | 1.00 (1.00, 1.00) | 0.63 |
| Hypertension | 0.86 (0.70, 1.05) | 0.14 |
| Diabetes | 1.48 (1.20, 1.82) | **<0.001** |
| Chronic pulmonary disease | 1.13 (0.90, 1.43) | 0.29 |
| Chronic kidney disease | 1.69 (1.37, 2.08) | **<0.001** |
| Hyperlipidemia | 1.01 (0.82, 1.25) | 0.92 |
| Coronary disease | 1.23 (0.99, 1.54) | 0.06 |
| Baseline ventilation status (within 48 hours of admission) |  |  |
| Room air | (reference) |  |
| Supplemental oxygen | 1.02 (1.00, 1.04) | **0.049** |
| Mechanical ventilation | 1.04 (1.02, 1.07) | **<0.001** |
| ABO blood group |  |  |
| A | 0.94 (0.75, 1.17) | 0.56 |
| B | 1.01 (0.75, 1.35) | 0.96 |
| AB | 1.21 (0.72, 2.01) | 0.47 |
| O | (reference) |  |
| Any steroids | 1.07 (1.05, 1.09) | **<0.001** |
| Hydroxychloroquine | 1.15 (0.73, 1.82) | 0.53 |
| Lopinavir/ritonavir | 0.89 (0.45, 1.77) | 0.74 |
| Remdesivir | 1.21 (0.94, 1.54) | 0.14 |
| Ribavirin | 1.35 (0.73, 2.51) | 0.34 |
| Tocilizumab | 1.03 (1.02, 1.04) | **<0.001** |
| HR: hazard ratio; CI: confidence interval | **C-statistic = 0.76** | |
| Baseline ventilation status, steroids, and tocilizumab were treated as time-varying covariates in this model | | |
