## Supplemental Table 4 for "Early transfusion of a large cohort of COVID-19 patients with high titer anti-SARS-CoV-2 spike protein IgG convalescent plasma confirms a signal of significantly decreased mortality"

**Table S4. Patient Characteristics in Secondary PS Matched Cohorts**

|  | **Secondary matched, all plasma titers** | | | | **Secondary matched, titer ≥1350** | | | |
| --- | --- | --- | --- | --- | --- | --- | --- | --- |
|  | **Total** | **Not Transfused** | **Transfused** | **P-value** | **Total** | **Not Transfused** | **Transfused** | **P-value** |
|  | **(N=935)** | **(*n*=594)** | **(*n*=341)** |  | **(N=903)** | **(*n*=582)** | **(*n*=321)** |  |
| Age (years) |  |  |  | 0.81 |  |  |  | 0.07 |
| <30 | 30 (3.2) | 19 (3.2) | 11 (3.2) |  | 36 (4.0) | 21 (3.6) | 15 (4.7) |  |
| 30-39 | 132 (14.1) | 80 (13.5) | 52 (15.2) |  | 114 (12.6) | 63 (10.8) | 51 (15.9) |  |
| 40-49 | 192 (20.5) | 118 (19.9) | 74 (21.7) |  | 175 (19.4) | 106 (18.2) | 69 (21.5) |  |
| 50-59 | 245 (26.2) | 155 (26.1) | 90 (26.4) |  | 242 (26.8) | 159 (27.3) | 83 (25.9) |  |
| 60-69 | 210 (22.5) | 134 (22.6) | 76 (22.3) |  | 212 (23.5) | 144 (24.7) | 68 (21.2) |  |
| 70-79 | 103 (11.0) | 72 (12.1) | 31 (9.1) |  | 89 (9.9) | 61 (10.5) | 28 (8.7) |  |
| ≥80 | 23 (2.5) | 16 (2.7) | 7 (2.1) |  | 35 (3.9) | 28 (4.8) | 7 (2.2) |  |
| Sex |  |  |  | 0.50 |  |  |  | 0.63 |
| Female | 414 (44.3) | 268 (45.1) | 146 (42.8) |  | 395 (43.7) | 258 (44.3) | 137 (42.7) |  |
| Male | 521 (55.7) | 326 (54.9) | 195 (57.2) |  | 508 (56.3) | 324 (55.7) | 184 (57.3) |  |
| Race |  |  |  | 0.053 |  |  |  | 0.07 |
| White | 605 (64.7) | 387 (65.2) | 218 (63.9) |  | 595 (65.9) | 383 (65.8) | 212 (66.0) |  |
| Black | 218 (23.3) | 149 (25.1) | 69 (20.2) |  | 201 (22.3) | 141 (24.2) | 60 (18.7) |  |
| Asian | 45 (4.8) | 22 (3.7) | 23 (6.7) |  | 46 (5.1) | 25 (4.3) | 21 (6.5) |  |
| Other | 32 (3.4) | 18 (3.0) | 14 (4.1) |  | 30 (3.3) | 18 (3.1) | 12 (3.7) |  |
| Unknown | 35 (3.7) | 18 (3.0) | 17 (5.0) |  | 31 (3.4) | 15 (2.6) | 16 (5.0) |  |
| Ethnicity |  |  |  | 0.64 |  |  |  | 0.20 |
| Non-Hispanic | 465 (49.7) | 302 (50.8) | 163 (47.8) |  | 447 (49.5) | 301 (51.7) | 146 (45.5) |  |
| Hispanic | 461 (49.3) | 286 (48.1) | 175 (51.3) |  | 448 (49.6) | 276 (47.4) | 172 (53.6) |  |
| Unknown | 9 (1.0) | 6 (1.0) | 3 (0.9) |  | 8 (0.9) | 5 (0.9) | 3 (0.9) |  |
| Body mass index (kg/m^2^), median (IQR) | 31.9 (27.6, 37.4) | 32.2 (27.8, 37.6) | 31.5 (27.5, 36.4) | 0.15 | 31.6 (27.6, 37.5) | 31.6 (27.5, 37.7) | 31.5 (27.7, 36.4) | 0.85 |
| Body mass index (kg/m^2^) |  |  |  | 0.65 |  |  |  | 0.46 |
| <18.5 | 3 (0.3) | 2 (0.3) | 1 (0.3) |  | 2 (0.2) | 1 (0.2) | 1 (0.3) |  |
| 18.5-24.9 | 98 (10.5) | 63 (10.6) | 35 (10.3) |  | 99 (11.0) | 70 (12.0) | 29 (9.0) |  |
| 25-29.9 | 254 (27.2) | 153 (25.8) | 101 (29.6) |  | 252 (27.9) | 156 (26.8) | 96 (29.9) |  |
| ≥30 | 580 (62.0) | 376 (63.3) | 204 (59.8) |  | 550 (60.9) | 355 (61.0) | 195 (60.7) |  |
| Body mass index ≥30 (kg/m^2^) |  |  |  | 0.29 |  |  |  | 0.94 |
| <30 | 355 (38.0) | 218 (36.7) | 137 (40.2) |  | 353 (39.1) | 227 (39.0) | 126 (39.3) |  |
| ≥30 | 580 (62.0) | 376 (63.3) | 204 (59.8) |  | 550 (60.9) | 355 (61.0) | 195 (60.7) |  |
| Body mass index ≥35 (kg/m^2^) |  |  |  | 0.15 |  |  |  | 0.60 |
| <35 | 614 (65.7) | 380 (64.0) | 234 (68.6) |  | 606 (67.1) | 387 (66.5) | 219 (68.2) |  |
| ≥35 | 321 (34.3) | 214 (36.0) | 107 (31.4) |  | 297 (32.9) | 195 (33.5) | 102 (31.8) |  |
| Body mass index ≥40 (kg/m^2^) |  |  |  | **0.047** |  |  |  | 0.43 |
| <40 | 782 (83.6) | 486 (81.8) | 296 (86.8) |  | 759 (84.1) | 485 (83.3) | 274 (85.4) |  |
| ≥40 | 153 (16.4) | 108 (18.2) | 45 (13.2) |  | 144 (15.9) | 97 (16.7) | 47 (14.6) |  |
| Hypertension |  |  |  | 0.30 |  |  |  | 0.28 |
| No | 448 (47.9) | 277 (46.6) | 171 (50.1) |  | 431 (47.7) | 270 (46.4) | 161 (50.2) |  |
| Yes | 487 (52.1) | 317 (53.4) | 170 (49.9) |  | 472 (52.3) | 312 (53.6) | 160 (49.8) |  |
| Diabetes |  |  |  | 0.43 |  |  |  | 0.28 |
| No | 541 (57.9) | 338 (56.9) | 203 (59.5) |  | 527 (58.4) | 332 (57.0) | 195 (60.7) |  |
| Yes | 394 (42.1) | 256 (43.1) | 138 (40.5) |  | 376 (41.6) | 250 (43.0) | 126 (39.3) |  |
| Chronic pulmonary disease |  |  |  | 0.37 |  |  |  | 0.47 |
| No | 819 (87.6) | 516 (86.9) | 303 (88.9) |  | 795 (88.0) | 509 (87.5) | 286 (89.1) |  |
| Yes | 116 (12.4) | 78 (13.1) | 38 (11.1) |  | 108 (12.0) | 73 (12.5) | 35 (10.9) |  |
| Chronic kidney disease |  |  |  | 0.27 |  |  |  | 0.06 |
| No | 810 (86.6) | 509 (85.7) | 301 (88.3) |  | 772 (85.5) | 488 (83.8) | 284 (88.5) |  |
| Yes | 125 (13.4) | 85 (14.3) | 40 (11.7) |  | 131 (14.5) | 94 (16.2) | 37 (11.5) |  |
| Hyperlipidemia |  |  |  | 0.41 |  |  |  | 0.12 |
| No | 626 (67.0) | 392 (66.0) | 234 (68.6) |  | 589 (65.2) | 369 (63.4) | 220 (68.5) |  |
| Yes | 309 (33.0) | 202 (34.0) | 107 (31.4) |  | 314 (34.8) | 213 (36.6) | 101 (31.5) |  |
| Coronary disease |  |  |  | 0.24 |  |  |  | 0.12 |
| No | 853 (91.2) | 537 (90.4) | 316 (92.7) |  | 820 (90.8) | 522 (89.7) | 298 (92.8) |  |
| Yes | 82 (8.8) | 57 (9.6) | 25 (7.3) |  | 83 (9.2) | 60 (10.3) | 23 (7.2) |  |
| Baseline ventilation status (within 48 hours of admission) |  |  |  | 0.04 |  |  |  | **0.02** |
| Room air | 49 (5.2) | 23 (3.9) | 26 (7.6) |  | 43 (4.8) | 19 (3.3) | 24 (7.5) |  |
| Supplemental oxygen | 832 (89.0) | 538 (90.6) | 294 (86.2) |  | 821 (90.9) | 539 (92.6) | 282 (87.9) |  |
| Mechanical ventilation | 54 (5.8) | 33 (5.6) | 21 (6.2) |  | 39 (4.3) | 24 (4.1) | 15 (4.7) |  |
| Ventilation status at Day 0 |  |  |  | 0.97 |  |  |  | 0.99 |
| Room air | 79 (8.4) | 52 (8.8) | 27 (7.9) |  | 81 (9.0) | 54 (9.3) | 27 (8.4) |  |
| Low flow | 566 (60.5) | 359 (60.4) | 207 (60.7) |  | 549 (60.8) | 353 (60.7) | 196 (61.1) |  |
| High flow/NIPPV | 238 (25.5) | 151 (25.4) | 87 (25.5) |  | 234 (25.9) | 149 (25.6) | 85 (26.5) |  |
| Mechanical ventilation | 48 (5.1) | 30 (5.1) | 18 (5.3) |  | 36 (4.0) | 24 (4.1) | 12 (3.7) |  |
| ECMO | 4 (0.4) | 2 (0.3) | 2 (0.6) |  | 3 (0.3) | 2 (0.3) | 1 (0.3) |  |
| ABO blood group |  |  |  | 0.34 |  |  |  | 0.72 |
| A | 234 (29.8) | 122 (27.4) | 112 (32.8) |  | 229 (30.5) | 126 (29.3) | 103 (32.1) |  |
| B | 109 (13.9) | 67 (15.1) | 42 (12.3) |  | 102 (13.6) | 63 (14.7) | 39 (12.1) |  |
| AB | 21 (2.7) | 13 (2.9) | 8 (2.3) |  | 16 (2.1) | 9 (2.1) | 7 (2.2) |  |
| O | 422 (53.7) | 243 (54.6) | 179 (52.5) |  | 404 (53.8) | 232 (54.0) | 172 (53.6) |  |
| Rh blood group |  |  |  | 0.29 |  |  |  | 0.27 |
| Negative | 71 (9.0) | 36 (8.1) | 35 (10.3) |  | 65 (8.7) | 33 (7.7) | 32 (10.0) |  |
| Positive | 715 (91.0) | 409 (91.9) | 306 (89.7) |  | 686 (91.3) | 397 (92.3) | 289 (90.0) |  |
| Interleukin-6 (pg/mL) at Day 0, median (IQR) | 57.0 (24.0, 127.0) | 52.5 (20.0, 125.0) | 63.5 (28.5, 133.0) | **0.03** | 53.0 (24.0, 123.0) | 52.0 (20.5, 122.5) | 59.0 (28.0, 123.5) | 0.17 |
| C-reactive protein (mg/dL) at Day 0, median (IQR) | 9.8 (5.3, 17.4) | 9.7 (5.1, 18.1) | 9.9 (5.5, 16.1) | 0.73 | 9.7 (5.3, 16.1) | 9.7 (5.2, 16.7) | 9.8 (5.5, 15.8) | 0.96 |
| Ferritin (ng/mL) at Day 0, median (IQR) | 847.0 (414.0, 1657.0) | 865.5 (379.0, 1777.0) | 814.0 (451.0, 1476.0) | 0.50 | 839.0 (436.0, 1637.0) | 889.0 (431.0, 1782.0) | 786.5 (438.0, 1454.0) | 0.08 |
| Fibrinogen (mg/dL) at Day 0, median (IQR) | 651.0 (554.5, 749.5) | 659.0 (569.0, 766.0) | 642.0 (526.0, 738.0) | 0.050 | 654.5 (556.0, 748.0) | 658.0 (574.0, 761.0) | 646.0 (530.0, 736.0) | 0.12 |
| D-dimer (µg/mL FEU) at Day 0, median (IQR) | 1.0 (0.6, 1.8) | 1.1 (0.6, 2.0) | 0.8 (0.6, 1.5) | **0.004** | 0.9 (0.6, 1.6) | 1.0 (0.6, 1.7) | 0.8 (0.6, 1.5) | 0.06 |
| Concomitant medication |  |  |  |  |  |  |  |  |
| Any steroids | 697 (74.5) | 452 (76.1) | 245 (71.8) | 0.15 | 663 (73.4) | 438 (75.3) | 225 (70.1) | 0.09 |
| Dexamethasone | 441 (47.2) | 293 (49.3) | 148 (43.4) | 0.08 | 441 (48.8) | 294 (50.5) | 147 (45.8) | 0.17 |
| Hydrocortisone | 68 (7.3) | 44 (7.4) | 24 (7.0) | 0.83 | 61 (6.8) | 42 (7.2) | 19 (5.9) | 0.46 |
| Methylprednisolone | 393 (42.0) | 249 (41.9) | 144 (42.2) | 0.93 | 364 (40.3) | 239 (41.1) | 125 (38.9) | 0.53 |
| Prednisone | 145 (15.5) | 95 (16.0) | 50 (14.7) | 0.59 | 132 (14.6) | 89 (15.3) | 43 (13.4) | 0.44 |
| Azithromycin | 704 (75.3) | 454 (76.4) | 250 (73.3) | 0.29 | 664 (73.5) | 432 (74.2) | 232 (72.3) | 0.52 |
| Hydroxychloroquine | 120 (12.8) | 74 (12.5) | 46 (13.5) | 0.65 | 92 (10.2) | 65 (11.2) | 27 (8.4) | 0.19 |
| Lopinavir/ritonavir | 18 (1.9) | 14 (2.4) | 4 (1.2) | 0.32 | 9 (1.0) | 7 (1.2) | 2 (0.6) | 0.50 |
| Remdesivir | 352 (37.6) | 230 (38.7) | 122 (35.8) | 0.37 | 340 (37.7) | 220 (37.8) | 120 (37.4) | 0.90 |
| Ribavirin | 62 (6.6) | 43 (7.2) | 19 (5.6) | 0.32 | 31 (3.4) | 23 (4.0) | 8 (2.5) | 0.25 |
| Tocilizumab | 452 (48.3) | 291 (49.0) | 161 (47.2) | 0.60 | 431 (47.7) | 283 (48.6) | 148 (46.1) | 0.47 |

Values are in median (interquartile range, IQR) for continuous variables and number (%) for categorical variables.

Difference between groups was compared using the Wilcoxon rank-sum test for continuous variables and Chi-square test of Fisher’s exact tests for categorical variables, as appropriate.

ECMO: extracorporeal membrane oxygenation; NIPPV: noninvasive positive pressure ventilation
